# The urban physical exposome and leisure-time physical activity in early midlife: a FinnTwin12 study

**DOI:** 10.1101/2024.06.09.24308658

**Authors:** Zhiyang Wang, Sari Aaltonen, Roos Teeuwen, Vasileios Milias, Carmen Peuters, Bruno Raimbault, Teemu Palviainen, Erin Lumpe, Danielle Dick, Jessica E. Salvatore, Maria Foraster, Payam Dadvand, Jordi Júlvez, Achilleas Psyllidis, Irene van Kamp, Jaakko Kaprio

**Author notes:** Corresponding author: Jaakko Kaprio; address: Institute for Molecular Medicine, University of Helsinki, PL 20 (Tukholmankatu 8), FI-00014, Helsinki, Finland.

## Abstract

Leisure-time physical activity is beneficial for health and is associated with various urban characteristics. Using the exposome framework, the totality of the environment, this study investigated how urban physical environments were associated with leisure-time physical activity during early midlife. A total of 394 participants (mean age: 37, range 34-40) were included from the FinnTwin12 cohort residing in five major Finnish cities in 2020. We comprehensively curated 145 urban physical exposures at residential addresses of participants and measured three leisure-time physical activity measures: (1) total leisure-time physical activity (total LTPA) and its sub-domains (2) leisure-time physical activity without commuting activity (LTPA) and (3) commuting activity. Using K-prototypes cluster analysis, we identified three urban clusters: “original city center,” “new city center,” and “suburban”. Results from adjusted linear regression models showed that participants in the “suburban” cluster had lower levels of total LTPA (beta: -0.13, 95% CI: -0.23, -0.03) and LTPA (beta: -0.17, 95% CI: -0.28, -0.05), compared to those in the “original city center” cluster. The eXtreme Gradient Boosting models ranked exposures related to greenspaces, pocket parks, and road junctions as the top important factors influencing outcomes, and their relationships with outcomes were largely non-linear. More road junctions and more pocket parks correlated with higher total LTPA and LTPA. When the all-year normalized difference vegetation index within a 500 m buffer fell below 0.4, it correlated with higher levels of total LTPA, whereas above 0.4, it correlated with lower levels. To conclude, our findings revealed a positive correlation between urbanicity and physical activity in Finnish cities and decomposed this complexity into crucial determinants. Importance rankings and nonlinear patterns offer valuable insights for future policies and projects targeting physical inactivity.

## 1 Introduction

Regular physical activity has been widely demonstrated to prevent multiple non-communicable diseases and reduce the risk of premature death^1^. The economic and health burden arising from physical inactivity is substantial and continually rising, costing public health care systems an estimated USD 47.6 billion globally every year^2^. Since previous studies show a strong contribution of environmental factors to physical activity^3–5^, interventions targeting the environment may be a good entry point to promote physical activity.

Urbanization stands as a transformative trend, with more than half the world’s population currently residing in urban areas^6^. Many reviews have summarized the salient link between the urban environment and physical activity^7–9^. The exposome offers a theoretical framework with an umbrella perspective to depict the totality of the environment that people experience^10^ and examines health effects from the real-world urban environment, of which the urban physical component plays an important role. The exposome studies have the potential to unveil more comprehensive non-genetic predictors through large-scale characterization of the environment. Gorman et al. have outlined the bidirectional effects between the exposome and physical activity but pointed out the uncertainty in mechanism and interactions^11^. The urban physical exposome is ubiquitous and multifaceted, which makes it a complex entity to study.

Every environmental factor contributes to this complex totality of exposures, and no factor is isolated. Urban regeneration projects are a good example, usually designed to improve public health by implementing structural and risk-minimizing solutions. They often yield collateral effects on other aspects, such as bringing economic, social, and cultural benefits, within the city’s complex system^12–14^. For example, an urban riverside park regeneration project in Barcelona, Spain was estimated to attract over five thousand adult users daily to perform different types of physical activity^15^. Beyond the project’s basic objectives, an open-air museum will be built there, transforming social and built environments. Nowadays, regeneration projects around the world are often multi-component and intersectoral. In another Barcelona regeneration program, aiming to improve living conditions in the most disadvantaged neighborhoods (involving, for example, social services, green spaces, and household support), researchers found that the neighborhood with a bigger project budget was associated with a higher frequency of physical activity among residents^16^. Previous studies relying on single exposures or limited sets were relatively inadequate to depict the broader urban environment and its health effects.

In this study, we aimed to comprehensively study the impact of the urban physical component of the exposome on the level of leisure-time physical activity during early midlife through two objectives: 1) clustering people with heterogeneous urban environments and comparing physical activity levels between clusters, as well as sex-specific effect and 2) ranking urban physical exposures by importance on leisure-time physical activity, examining non-linear relationships, and detecting pairwise interactions between exposures.

## 2 Material and methods

The flow chart of this study is presented in Figure 1.

**Figure 1.**
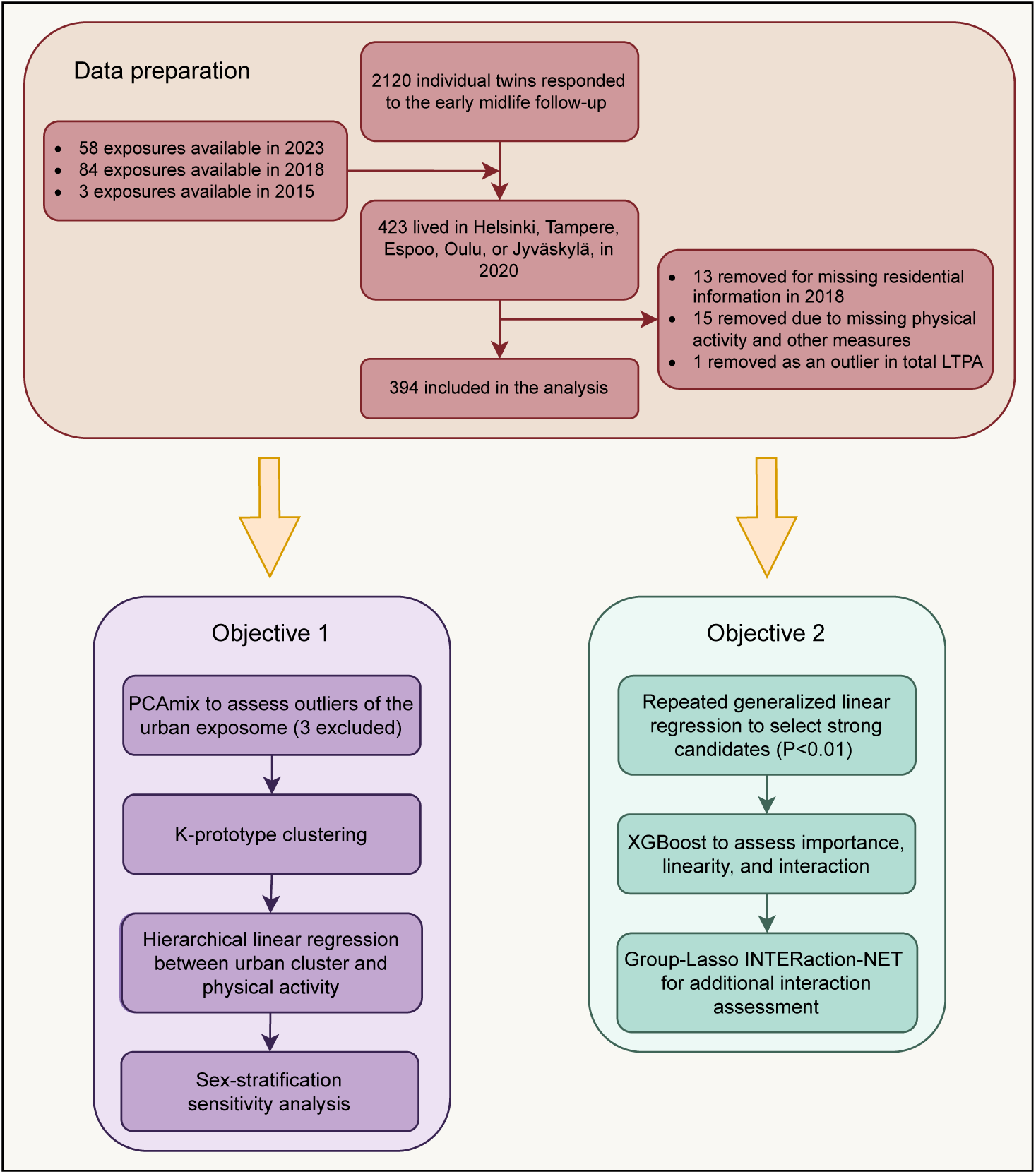
Study flow

### 2.1 Participants

Participants were from the FinnTwin12 cohort, which is a nationwide prospective cohort of all Finnish twins born between 1983 and 1987. Briefly, at baseline (1994-1999), 5522 12-year-old twins were invited to participate and 87% of them agreed to take part. There were four follow-ups: age 14, age 17, young adulthood (mean age 24), and early midlife (mean age 37), with retention rates of 92%, 75%, 66%, and 41%, respectively. A recent study has detailed the latest follow-up of the cohort^17^. In this study, we included individual twins who lived in five large cities of Finland, namely Helsinki, Tampere, Espoo, Oulu, or Jyväskylä, in 2020. A

### 2.2 Measures

#### 2.2.1 Leisure-time physical activity

Our study focuses on early midlife leisure-time physical activity, which is performed at the person’s discretion along with essential daily living activity or work-related tasks^18^. This type of physical activity is considered one of the most effective ways to increase overall physical activity levels^19^. It was measured through structured and validated questions on the frequency, mean duration, and mean intensity of participants’ leisure-time physical activity sessions, as well as a question on their commuting activity^20,21^. Based on these structured questions, we quantified mean metabolic equivalent of task (MET) hours per day, which expresses the energy cost of physical activities in the form of the resting metabolic rate^22^. Its calculation formula was the following: physical activity frequency × physical activity duration × physical activity intensity^23^. The MET values for activity intensity were: 4 for intensity corresponding to walking, 6 for intensity corresponding to vigorous walking to jogging, 10 for intensity corresponding to jogging, and 13 for the intensity corresponding to running. All types of leisure time physical activities were considered when MET hours per day were calculated. We assumed that commuting activity was done on 5 days per week and on the intensity of walking. The questions are listed in Supplemental Note 1.

The primary measure, *total leisure-time physical activity (total LTPA)*, was the sum of leisure-time and commuting-related physical activities. These two sub-domains were secondary measures: 1) *leisure-time physical activity without commuting activity (LTPA)* and 2) *commuting activity*. Participants with over mean 45 MET hours/day of total LTPA were identified as outliers and removed. This threshold corresponds to, for example, approximately 3.5 hours of fast running daily, which is likely unrealistic^24^. The distributions of all three measures are shown in Supplemental Figure 1, and due to the skewness, we log-transformed them.

#### 2.2.2 Urban physical exposome

We assigned 145 indicators of urban physical exposures to the residential address of each study participant. Detailed description and summary statistics of these indicators are presented in Supplemental Table 1. The urban physical exposome set comprehensively depicted the urban environment including aspects such as traffic, streets, land use, green (i.e. parks, forests, and fields) and blue (i.e. lakes and seas) spaces, and so on. The computing and enriching process was on the geocode level and derived from multiple open sources, which is described in Supplemental Note 2 and elsewhere^25–28^. Most urban physical exposures were measured or modelled in 2018 and 2023, and the percentage of area covered by trees was measured in 2015. We used the residential history provided by the Digital and Population Data Services Agency, Finland between birth and 2021 to merge the urban physical exposures by EUREF-FIN geocodes. Exposures available in 2018 or 2015 were merged with residential addresses of participants in 2018 or 2015, while exposures available in 2023 were merged with residential addresses in 2020.

#### 2.2.3 Other measures

Five sociodemographic variables were identified a priori: sex (categorical, female vs. male), age (continuous, year), work (categorical, not working or other situation vs. currently work), education (categorical, post-secondary or lower vs. bachelor/equivalent or above), and marital status (categorical, married, steady relationship, or living together vs. no). The latter three were self-reported at the early midlife follow-up. Sex was based on the register information obtained when the cohort was established, while age was computed from the difference between the date of response and the date of birth. There were another three behavioral variables: illicit substance use (categorical, never vs. at least once), ever smoker (smoked over 100 cigarettes lifetime) (categorical, no vs. yes), and alcohol drinking (categorical, monthly or less or even never vs. 2-4 times a month or more), inquired also at the early midlife follow-up. Adult leisure-time physical activity was associated to most of the sociodemographic and behavioral variables, as shown in previous research^29–31^.

To depict the social environment, four neighborhood social variables at the postal code level were derived from Statistics Finland in 2018: the proportion of resident living alone (single household), of residents with the lowest education level, of residents with the lowest income quartile, and of unemployed residents. A neighborhood deprivation score was generated from the latter three social variables^32^. We first standardized the three variables to z-scores, and their mean value is the deprivation score. Using a median split, we then categorized neighborhoods where participants lived in 2018 into two levels: low- and high-deprived. Thus, there were two neighborhood social variables: the proportion of resident living alone and deprivation level, which were merged via residential history in 2018 too.

### 2.3 Analysis

#### 2.3.1 Data processing

After excluding those people who did not have information on leisure-time physical activity, sociodemographic, behavioral, and neighborhood-level social variables, 394 twin individuals resident in these urban areas were included in this study. Given that there were only 44 twin pairs with both cotwins satisfying the inclusion criteria, we did not consider zygosity as a covariate and did not perform any pairwise twin analysis. The distribution of sociodemographic and behavioral variables among included and excluded participants are presented in Supplemental Table 2, respectively. There were significant differences between included and excluded participants in education, illicit substance use, and alcohol drinking.

#### 2.3.2 Clustering analysis

The k-prototypes cluster analysis was employed to distinguish distinct patterns in the urban environment. It combines dissimilarity measures from both k-means and -modes algorithms for mixed types of exposures, and has shown to have a good performance^33,34^. Continuous exposures were standardized by standard deviation (SD). All 145 urban physical exposures were included in the clustering algorithms. The Silhouette method was used to pre-specify the number of clusters^35^. One-step imputation within the algorithm was applied for missing values^36^. Since k-prototypes cluster analysis is sensitive to outliers, the principal component analysis of mixed data was conducted before. Participants whose first or second principal components fell outside the range of five standard deviations were identified as outliers^37^, as a practical way, and excluded from the cluster analysis; three participants were excluded.

Next, hierarchical linear regression was performed for the relationship between the urban cluster and leisure-time physical activity measures with three adjustment plans for covariates: 1) sociodemographic variables, 2) sociodemographic and behavioral variables, and 3) sociodemographic, behavioral, and neighborhood social variables. The cluster effect of sampling based on families of twin pairs was controlled by the robust standard error. We also performed the sex-stratified analysis.

#### 2.3.3 Machine learning analysis

Before exploring the complexity within the urban environment via a pluralistic analysis platform, generalized linear regression models with the robust standard error were repeatedly performed between each leisure-time physical activity measure (total LTPA, LTPA, and commuting activity) and each urban physical exposure (missing values were imputed). The *a priori* significant threshold of 0.01 was used to identify noteworthy candidates. Dimensional reduction increases the model stability of subsequent analysis.

Then, we performed the eXtreme Gradient Boosting (XGBoost) model to assess the importance of urban physical exposures on each physical activity measure, uncover interactions, and identify nonlinear relationships^38^. It is an optimized distributed gradient boosting library designed for efficient and scalable training of machine learning models, with gradient-boosted decision trees algorithm^38^. The hyperparameters were tuned through the 5-fold cross-validation grid search^39^. The participants were randomly split into training and testing subsets in a ratio of 3:1. The model performance was evaluated by root-mean-square error (RMSE). All urban physical exposures, sociodemographic, behavioral, and neighborhood social variables were included in the model. After hyperparameter tuning, the model was repeated two additional times with different seeds for result robustness.

To increase model transparency, the SHapley Additive exPlanations (SHAP) value was used to interpret and visualize the results from the XGboost model, which features the exposures’ importance on the outcome based on the cooperative game theory^40^. Its direction suggests the direction of impact on prediction, leading the model to predict either a higher or lower value of outcomes. Its magnitude is a measure of how strong the effect is. We quantified pairwise interaction SHAP values between included variables and summed their absolute value of all participants, with a high value indicating a strong interaction and synergistic effect^41^. Additionally, Group-Lasso INTERaction-NET was performed for interaction to compare with the XGBoost’s result^42^.

#### 2.3.4 Sensitivity analysis

Due to missing values in urban physical exposures, we additionally performed sensitivity analyses of K-prototype cluster analysis and repeated generalized linear regression models between each urban physical exposure and each leisure-time physical activity measure, after removing participants with missing values (n=13).

## 3 Results

### 3.1 Description of participants

Of the 394 included participants (mean age: 37, SD: 1.5) (Table 1), more individuals were female (55%). Altogether, 87%, 79%, and 75% of participants were employed, had at least bachelor-level education, and were married or in a stable relationship, respectively. In their early midlife, more than half of the participants drank alcohol at least 2-4 times a month (58%), but fewer had smoked over 100 cigarettes (45%) or had used illicit substances such as marijuana at least once (48%). Before log-transformation, the means of total LTPA, LTPA, and commuting activity (unit: MET hours/day) were 5.4 (SD: 4.7), 4.3 (SD: 4.4), and 1.1 (SD: 1.0), respectively. After log-transformation, Spearman correlations between total LTPA and LTPA, between total LTPA and commuting activity, and between LTPA and commuting activity were 0.9, 0.3, and 0.1, respectively

**Table 1:** Characteristics of sociodemographic, behavior, and neighborhood social variables (participants n=394)

| **Characteristics** | **N. (%) / Mean (SD)** |
| --- | --- |
| **Sex** |  |
| Male | 179 (45.4) |
| Female | 215 (54.6) |
| **Work** |  |
| Not working or other situation | 51 (12.9) |
| Currently work | 343 (87.1) |
| **Education** |  |
| Post-secondary or lower | 84 (21.3) |
| Bachelor/equivalent or above | 310 (78.7) |
| **Marital status** |  |
| No | 97 (24.6) |
| Married, steady relationship, or<br>living together | 297 (75.4) |
| **Age (years)** | 37.1 (1.5) |
| **Illicit substance use** |  |
| No | 204 (51.8) |
| Yes | 190 (48.2) |
| **Ever smoker (smoked over 100**<br>**cigarettes lifetime)** |  |
| No | 216 (54.8) |
| Yes | 178 (45.2) |
| **Alcohol** |  |
| Monthly or less, or even never | 164 (41.6) |
| 2-4 times a month or more | 230 (58.4) |
| **Deprivation level** |  |
| Low | 225 (57.1) |
| High | 169 (42.9) |
| **The proportion of single**<br>**households in the neighborhood** | 50.0 (10.8) |

### 3.2 Results from clustering and hierarchical regression

The Silhouette method identified the optimal number of clusters to be three (largest Silhouette index, total within-cluster sum of squares: 42323.69). Using the map of Helsinki and Espoo and the spatial layer of centers and shopping areas in 2019 from the community structure monitoring system, Finnish Environment Institute^43^, we visually classified Cluster 1, 2, and 3 as the “Original city center”, “New city center”, and “Suburban” clusters, respectively, based on the participants’ residence in 2018, as the urban cluster variable (Figure 2).

**Figure 2:**
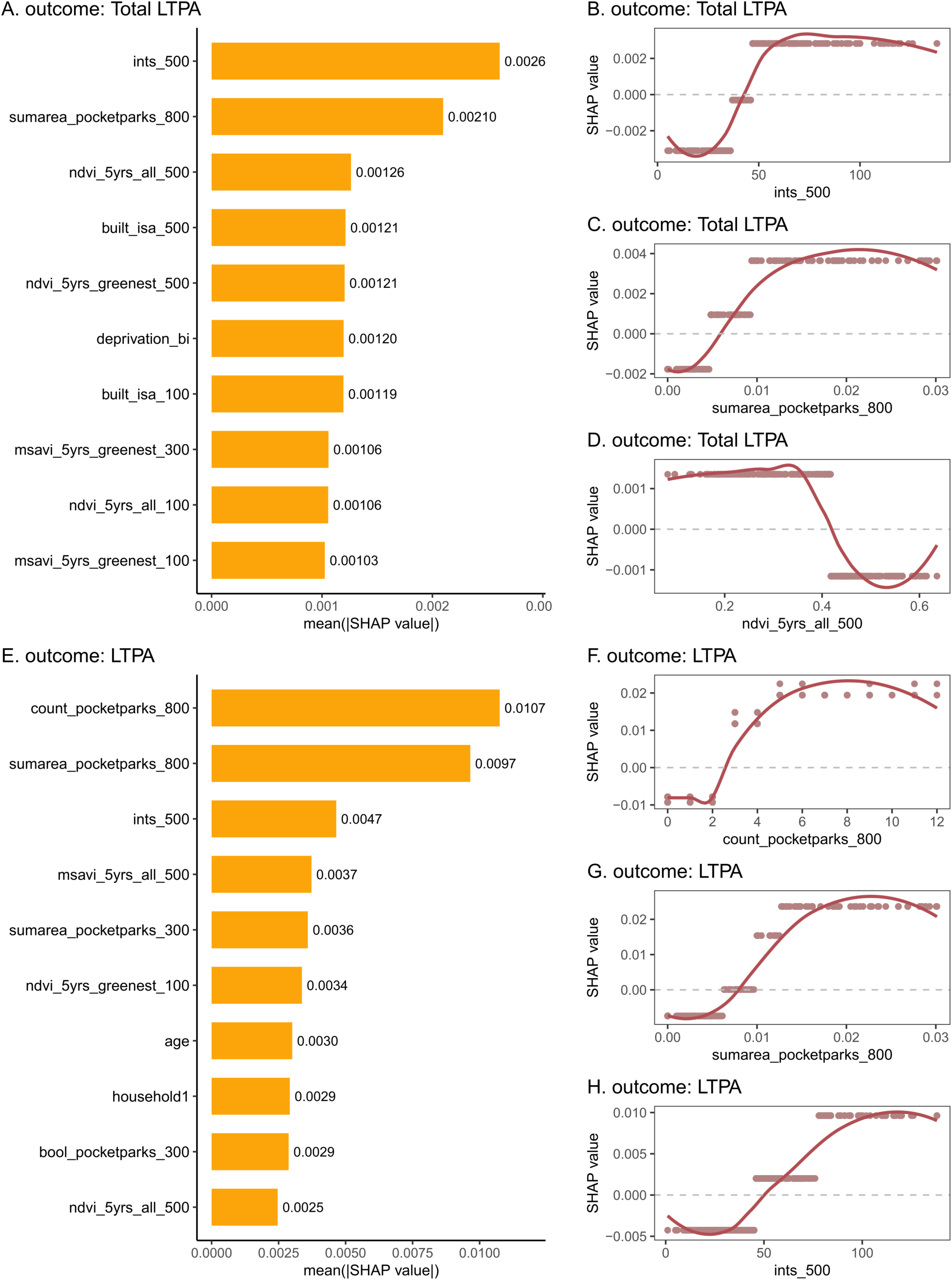
Twin participants’ residence in the Helsinki and Espoo area in 2018 colored by cluster. Note: The gray layer shows centers and shopping areas in 2019.

After fully adjusting for sociodemographic, behavioral, and neighborhood social variables, compared to participants who lived in the “original city center” cluster, participants who lived in the “suburban” cluster were associated with significantly lower log-transformed scores of total LTPA (beta: -0.13, 95% CI: -0.23, - 0.03) and LTPA (beta: -0.17, 95% CI: -0.28, -0.05) (Table 2). The effect sizes did not change substantially after adjustment of sociodemographic variables only and adjustment of both sociodemographic and behavioral variables. Regardless of adjustment plans, there was no significant association between the urban cluster and commuting activity (Table 2). There was no significant difference in any outcome between participants who lived in the “suburban” and “new city center” clusters. The powers of full-adjusted models of total LTPA, LTPA, and commuting activity were all 1.0.

**Table 2:**
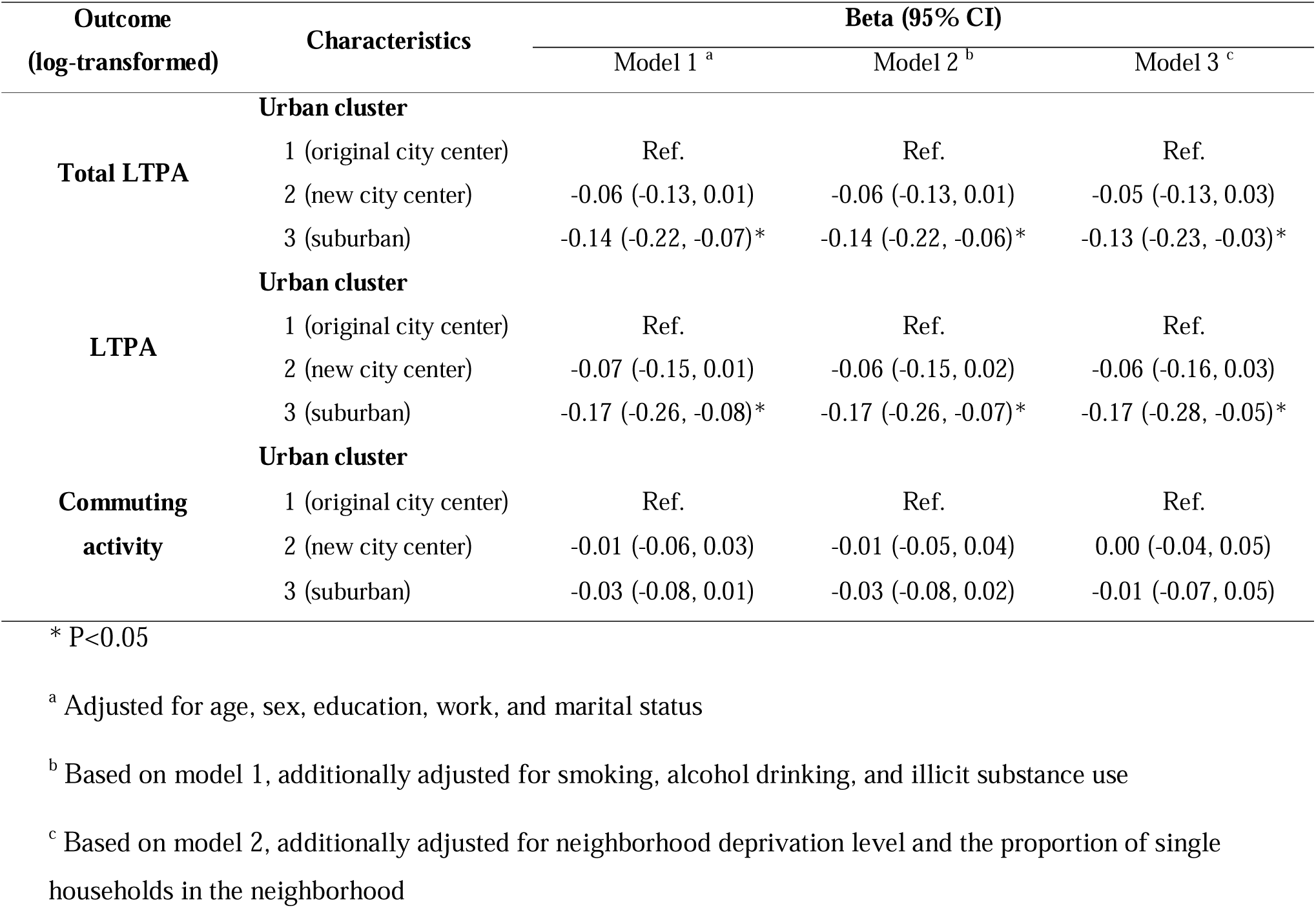
Results of the linear regression between the urban cluster and physical activity measures.

| Outcome<br>(log-transformed) | Characteristics | Beta (95% CI) |  |  |
| --- | --- | --- | --- | --- |
|  |  | Model 1 <sup>a</sup> | Model 2 <sup>b</sup> | Model 3 <sup>c</sup> |
| **Total LTPA** | **Urban cluster** |  |  |  |
|  | 1 (original city center) | Ref. | Ref. | Ref. |
|  | 2 (new city center) | -0.06 (-0.13, 0.01) | -0.06 (-0.13, 0.01) | -0.05 (-0.13, 0.03) |
|  | 3 (suburban) | -0.14 (-0.22, -0.07)* | -0.14 (-0.22, -0.06)* | -0.13 (-0.23, -0.03)* |
| **LTPA** | **Urban cluster** |  |  |  |
|  | 1 (original city center) | Ref. | Ref. | Ref. |
|  | 2 (new city center) | -0.07 (-0.15, 0.01) | -0.06 (-0.15, 0.02) | -0.06 (-0.16, 0.03) |
|  | 3 (suburban) | -0.17 (-0.26, -0.08)* | -0.17 (-0.26, -0.07)* | -0.17 (-0.28, -0.05)* |
| **Commuting**<br>**activity** | **Urban cluster** |  |  |  |
|  | 1 (original city center) | Ref. | Ref. | Ref. |
|  | 2 (new city center) | -0.01 (-0.06, 0.03) | -0.01 (-0.05, 0.04) | 0.00 (-0.04, 0.05) |
|  | 3 (suburban) | -0.03 (-0.08, 0.01) | -0.03 (-0.08, 0.02) | -0.01 (-0.07, 0.05) |

\* P<0.05
<sup>a</sup> Adjusted for age, sex, education, work, and marital status
<sup>b</sup> Based on model 1, additionally adjusted for smoking, alcohol drinking, and illicit substance use
<sup>c</sup> Based on model 2, additionally adjusted for neighborhood deprivation level and the proportion of single households in the neighborhood

After stratifying the analyses based on sex, we observed that, in males, the result pattern and effect sizes were like the overall results between the urban cluster and total LTPA, while the association between the urban cluster and LTPA became null after full adjustment (Supplemental Table 3). However, in females, after additionally adjusting for behavioral variables only or for both behavioral and neighborhood social variables, no significant association of the urban cluster with total LTPA and LTPA was seen (Supplemental Table 3).

### 3.3 Results from XGBoost

Based on the repeated generalized linear regression, there were 25 urban physical exposures significantly associated with total LTPA and 24 with LTPA (Supplemental Table 4). No urban physical exposure met the threshold p-value of 0.01 for association with commuting activity (Supplemental Table 4), so there was no XGBoost analysis for it.

In the XGBoost model of total LTPA including all urban physical exposures, sociodemographic, behavioral, and neighborhood social variables, the top three important urban physical exposures were the count of any type of road junctions within a 500 m buffer (ints_500), the total area of all interconnected pocket parks within an 800 m walking distance (sumarea_pocketparks_800), and the 5-years moving average of Normalized Difference Vegetation Index (NDVI), an indicator of general greenness, within a 500 m buffer around the home during whole year (ndvi_5yrs_all_500) (Figure 3A). In dependence plots, SHAP values positively correlated with both the count of any type of road junctions within a 500 m buffer (Figure 3B) and the total area of all interconnected pocket parks within an 800 m walking distance (Figure 3C). When the two urban physical exposures were within a certain range, SHAP values remained constant, which this type of non-linearity made these two exposures look like threshold variables. When the count of any type of road junctions within a 500 m buffer was in the range of 1-40, 40-50, and over 50, the SHAP values were approximately -0.003, 0, and 0.003, respectively. When the total area of all interconnected pocket parks within an 800 m walking distance was in the range of 0-0.005, 0.005-0.01, and over 0.01 km^2^, the SHAP values were approximately -0.002, 0.001, and 0.004, respectively. In Figure 3D, the 5-years moving average of NDVI within a 500 m buffer during whole year also showed a pattern of a binary threshold variable. When it was below 0.4, the SHAP value was around 0.0012. When it was over 0.4, the SHAP value was around -0.0012.

**Figure 3:**
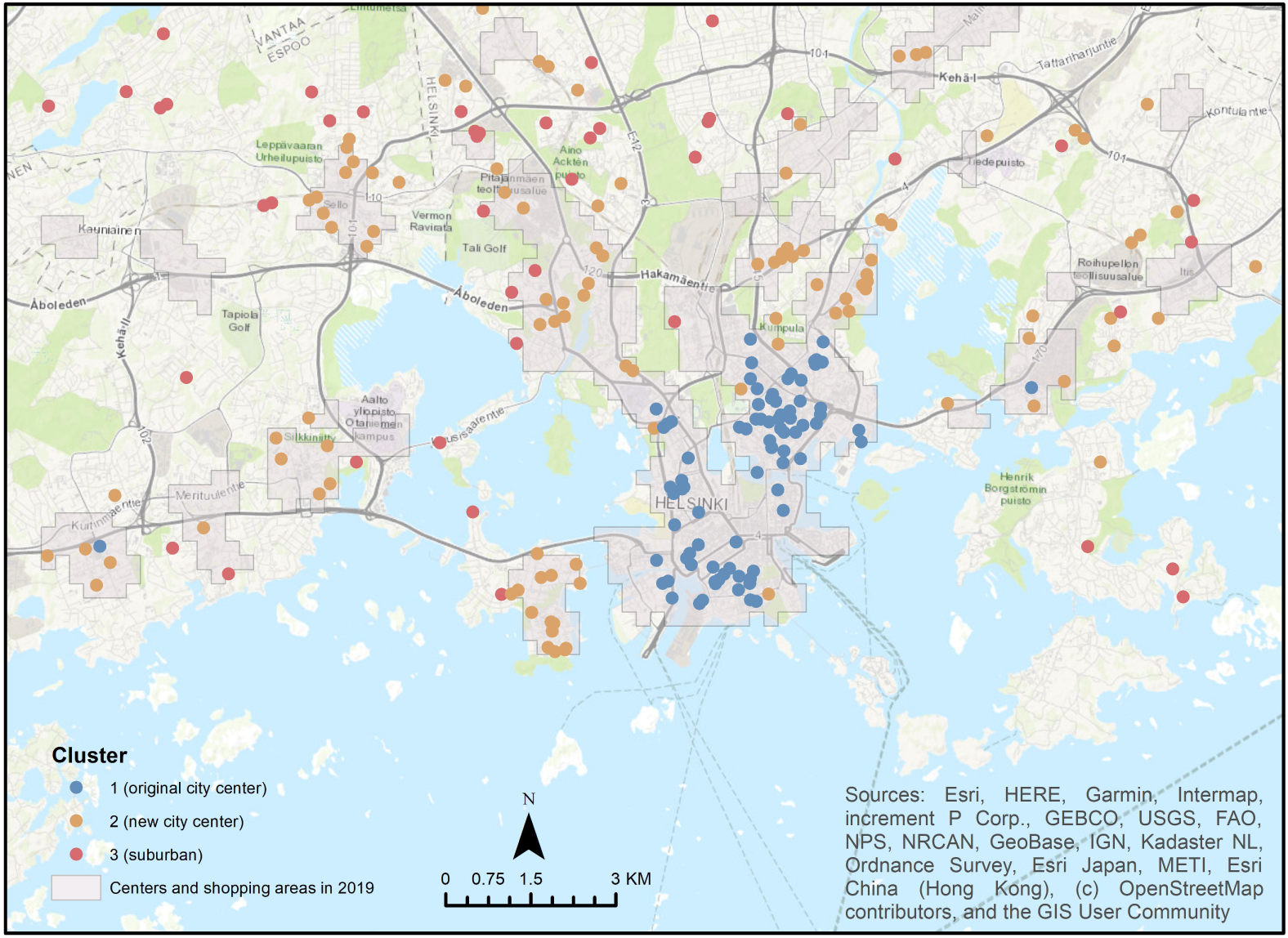
Results of XGBoost models for total leisure-time physical activity (total LTPA) and leisure-time physical activity without commuting activity (LTPA) Note: The SHAP bar plots show the influence of each variable: total LTPA (a) and LTPA (e). The SHAP dependence plots show how a single individual influences the XGboost prediction on total LTPA (b, c, d) and LTPA (f, g, h). ints_500 is the count of any type of road junctions within a 500 m buffer;sumarea_pocketparks_800 is the total area of all interconnected pocket parks within an 800 m walking distance; ndvi_5yrs_all_500 is the 5-years moving average of Normalized Difference Vegetation Index within a 500 m buffer during whole year; count_pocketparks_800 is the count of pocket parks within an 800 m walking distance. Abbreviation: leisure-time physical activity (LTPA); SHapley Additive exPlanations (SHAP)

In the XGBoost model of LTPA (Figure 3E), the most important urban physical exposures were the count of pocket parks within an 800 m walking distance (count_pocketparks_800), the total area of all interconnected pocket parks within an 800 m walking distance remained the second most important, and the count of any type of road junctions within a 500 m buffer became the third most important. The SHAP value revealed a switch in predictions from lower (negative SHAP values) to higher (positive SHAP values) log-transformed LTPA when the number (count) of pocket parks within an 800 m walking distance was more than two (Figure 3F). Interestingly, each count led to two different but close SHAP values. The patterns of the total area of all interconnected pocket parks within an 800 m walking distance (Figure 3G) and the count of any type of road junctions within a 500 m buffer (Figure 3H) were similar to the model of total LTPA.

Supplemental Figure 2 displays pairwise SHAP interaction values in the XGBoost model of total LTPA, and there was no pairwise interaction between urban physical exposures. Similarly, the XGBoost model of LTPA (Supplemental Figure 3) indicates slightly some interactions but with very low values (<0.001). Group-Lasso INTERaction-NET models also did not capture any strong pairwise interaction for the two physical activity measure analyses.

For the XGBoost model of total LTPA, the RMSE is 0.27 in the training subset and 0.29 in the testing subset. Comparing the reported test with two extra tests, the importance rank varied, but the count of any type of road junctions within a 500 m buffer was always the most or second most important (Supplemental Table 5). For the XGBoost model of LTPA, the RMSE is 0.32 in both training and testing subsets. The importance rank also varied between reported results and two extra tests, but the most important urban physical exposure was the count of pocket parks within an 800 m walking distance, being always in the top two (Supplemental Table 6).

### 3.4 Sensitivity analysis for missing value in urban physical exposures

After excluding 13 participants with missing values in some urban physical exposures, the Silhouette method identified two clusters. In the following fully adjusted linear regression models, no significant differences in any of the physical activity measures were found between the clusters. Repeated generalized linear regression analyses revealed 25 urban physical exposures significantly associated with total LTPA, consistent with the analysis using imputed data, and 26 exposures with LTPA, two more than the analysis with imputed data. Still, no urban physical exposure reached the 0.01 P-value threshold for association with commuting activity.

## 4 Discussion

We used clustering analysis and XGBoost to simultaneously and comprehensively study the effect of 145 urban physical exposures on leisure-time physical activity in 394 Finnish adults in their early midlife. Three clusters were identified: “original city center”, “new city center”, and “suburban”. We found people living in suburban areas had a lower level of physical activity in leisure time compared to those living in the original city center. There was no difference between “original city center” and “new city center” clusters. The effects appeared more clearly in males, while behavioral and neighborhood social factors may account for the associations in females. XGBoost models revealed a complex relationship between the urban physical exposome and leisure-time physical activities, in which important exposures showed non-linearity and looked like threshold variables. Increased road junctions and more and bigger pocket parks correlated with higher levels of leisure-time physical activity. However, higher amounts of vegetation greenness (indicated by NDVI) were associated with low leisure-time physical activity levels. We did not find any considerable interaction between urban physical exposures contributing to leisure-time physical activities.

Previous research has documented the relationship between the degree of urbanization and physical activity but with inconsistent findings regarding the direction of effects. A cross-sectional study in Shanghai, China with 327 respondents (mean age: 40) similarly reported higher leisure-time physical activity among downtown residents compared to suburban dwellers, but in contrast to our results, significant results were also found for transportation activities^44^. Another Canadian study showed that the physical activity level was higher in urban than in suburban among adolescents from schools in lower socio-economic areas^45^. Nevertheless, a systematic review suggested that children and teenagers who live in suburban areas were more physically active than in rural and urban areas^46^, and, similar to the Shanghai study above, a nationwide study in China showed that rising urbanization correlates with longer commuting times among adults (mean age: 45)^47^. Sex-specific effects have also been also observed. In the US, only male adolescents living in urban areas engaged in more moderate-to-vigorous physical activity (MVPA) than those living in suburban areas^48^. Additionally, distinct patterns between sexes in the significance and direction of associations between urbanity level in different aspects and physical activity measures were noted in Mexico^49^. Socioeconomic status (SES) might explain the sex difference, as the association weakened to null in females after adjusting for behavioral and neighborhood social variables. Previous population studies have observed some interaction effects between sex and SES on physical activity^50–52^. The inconsistency between literature and our findings may be due to different population characteristics, sports cultures, country contexts, urban planning, or urbanicity definitions. Instead of a pre-definition of (sub)urban areas by governmental guidelines, we used an unsupervised data-driven clustering method to determine heterogeneous urban environments within urban areas reflecting real-life exposure modes and accounting for correlation, additive, and mixture effects^53^.

XGBoost models ranked the elements of pocket parks, road junctions, and greenspaces as strongly associated with leisure-time physical activities among early midlife adults. A natural experimental study in low-income American neighborhoods found increased leisure-time exercise among middle-aged residents after pocket parks were constructed^54^. Users of pocket parks, defined as living within a 0.5 mile (∼800 m) radius, had higher exercise levels than traditional park users^54^. Researchers further summarized that pocket parks were cost-effective for promoting physical activity in inner-city areas^54^. A study in Chongqing, China, utilizing interviews on conceptual understanding of park images, revealed that the environmental characteristics of pocket parks contributed to a restorative effect involving entertainment activities and relief^55^. Noteworthy, a recent Chinese study using Light Gradient-Boosting Machine model found that recreational facilities were the most important factor for walking behavior in old adults but the number of parks was the least important among 11 factors, highlighting the specific effect driven by the content inside parks or recreation areas^56^. For road junctions, a Finnish study found the density of intersections, defined as the junction of a minimum of three roads, was positively associated with the number of physical activity bouts and the level of moderate to vigorous physical activity among older adults^57^. Zang et al. used random forest models to identify the intersection density, as well as streetscape greenery, as the most important physical exposure contributing to light physical activity among older adults^58^. More intersections usually indicate a greater degree of connectivity, which creates a more convenient environment for people to walk or bike to their destinations. However, the relationship between street connectivity, involving the number of intersections, and physical activity in all age groups of adults varied across different buffer areas in urban environments, suggesting the complexity of urban living environments^59^. Where the association of greenspace with physical activity is relatively inconsistent^60^, our findings show an association in which surrounding greenness is positively associated with LTPA up until a threshold of 0.4 NDVI, with higher NDVI relating to lower LTPA. High levels of green space might reflect suburban living to some extent, and other greenspace indicators, such as accessibility, were not prominent. The relationship between greenspace and physical activity could be moderated by the level of urbanization^60^. Other studies have similar findings on the threshold effect. For example, the positive association of physical activity with multiple green space uses indicators reached to peak when indicators were within a 600 m buffer^61^. Zang et al. also found that streetscape greenery had a positive effect on light physical activity when it ranged from 0.12 to 0.15 point, corresponding to a low level of visible greenery^58^. Besides, another Chinese study also identified the 0.4 NDVI, corresponding to areas with sparse to moderate vegetation, as the turning point for its association with self-rated health among the old population^62^, and self-rated health closely correlated with physical activity^63^. This annotation added to current evidence has critical guidance on urban planning.

We selected different methods to depict the contour of association between the urban physical exposome and leisure-time physical activity, translating abstract characteristics into practical understanding. On one hand, clustering analysis has the advantage of providing insight into real-world scenarios and holding a high scalability to uncover hidden patterns. On the other hand, tree-based machine learning can be applied as a pluralistic analysis platform to synthesize evidence between a range of urban physical exposures and physical activity^58,64,65^. Comparing to conventional analyses, the XGBoost model enhances our assessments with several advantages: 1) unraveling nonlinear relationships through visualization, 2) disentangling complex interactions among multiple exposures, and 3) offering robust computation for multi-inference approaches^66^. By deepening the understanding of distinct and complex characteristics of the urban physical exposome, supported by detailed exposure profiling, policymakers can develop precise and cost-effective interventions and strategies to address the challenge of low physical activity levels.

Besides its strength, this study is not without limitations. First, the sample size was relatively small compared to other exposome studies. Although the sample size for K-prototype clustering (over 10 times the number of clusters) and subsequent regression seems to be adequate (but not for the sex-stratified analysis), inconsistency in additional tests highlights the need for a larger sample. Additionally, due to the complexity of the large-dimensional exposome set, the modest sample size made capturing relatively small interactions more challenging. Therefore, we should be cautious when interpreting results. Second, only participants from the five largest cities in Finland were included, limiting the generalizability. Besides, we did not include any participants living in rural areas. Not only the physical environment, but lifestyles may also differ between urban and rural areas. Therefore, the interpretation should be narrowed down to specific types of cities. Third, urban physical exposures were based on residential addresses, which overlook dynamic human behaviors outside the home, leading to measurement errors. In addition, the used residential geocodes corresponded to participants’ residences in 2017, 2018, or 2020, without accounting for how long they lived at those addresses. Measurement errors could skew our identification of key determinants, as exposures with larger errors might show weaker associations and be classified as less influential, even if they are actually more important than those identified as most influential. More granular and accurate estimations of exposure and behavior could facilitate the exploration in the dynamic interaction between the environment and human behavior^12^. Fourth, some exposures were available in 2023 but merged with the address in 2020, posing a temporality issue. The relatively slow urban renewal and construction in Finland reduced the concern^67^. Fifth, missing values in exposures may introduce bias. Excluding participants with missing values altered the optimal number of clusters, while the number of significant associations between exposures and outcomes remained similar to the number based on imputed data. Given that only about 3% of participants had missing values, the effect is likely modest, but caution is still warranted. Sixth, leisure-time physical activity was self-reported. The device-based measurement of leisure-time physical activity would have been more accurate. However, the validity of leisure-time physical activity questions used in Finnish twins has been demonstrated^20,21^.

## 5 Conclusion

This study employed two analytical approaches to explore the intricate impact of the urban physical exposome on leisure-time physical activity in early midlife in Finland. Clustering analysis revealed three heterogeneous patterns of urban environments. Living in suburban areas was associated with lower levels of leisure-time physical activity than in original city center areas. XGBoost models identified pocket parks, road junctions, and greenspaces as influential factors with non-linear relationships, which behaved like threshold variables. Given limitations in sample size, generalizability, and measurement granularity, we call for further studies in other settings to replicate our analyses. We still advocate presenting the evidence to stakeholders and policymakers to develop tailored interventions on some urban features to achieve higher cost-effectiveness by focusing on the most influential determinants and their optimal ranges in addressing the challenge of the physically inactive lifestyle in our rapidly urbanizing world.

## Data Availability

The FinnTwin12 data are not publicly available due to the restrictions of informed consent. However, the FinnTwin12 data are available through the Institute for Molecular Medicine Finland (FIMM) Data Access Committee (DAC) for authorized researchers who have IRB/ethics approval and an institutionally approved study plan. To ensure the protection of privacy and compliance with national data protection legislation, a data use/transfer agreement is needed, the content and specific clauses of which will depend on the nature of the requested data. Requests will be addressed in a reasonable time frame (generally two to three weeks), and the primary mode of data access is by either personal visit or remote access to a secure server.

## Acknowledgments

This research was partly funded by the European Union’s Horizon 2020 research and innovation program under grant agreement No 874724 (Equal-Life). Equal-Life is part of the European Human Exposome Network. Data collection in FinnTwin12 has been supported by the National Institute on Alcohol Abuse and Alcoholism (grants AA-12502, AA-00145, and AA-09203 to Richard J. Rose, and AA015416 to Danielle Dick and Jessica Salvatore) and the Academy of Finland (grants 100499, 205585, 118555, 141054, 264146, 308248, 312073, 336823, and 352792 to Jaakko Kaprio). Jaakko Kaprio acknowledges support by the Academy of Finland (grants 265240, 263278). ISGlobal acknowledges support from the grant CEX2018-000806-S funded by MCIN/AEI/10.13039/501100011033, and support from the Generalitat de Catalunya through the CERCA Program.

## Author’s contribution

M.F., P.D., J.J., A.P., I.v.K., and J.K. conceived the exposome framework. Z.W. developed the research question and designed the analysis and other authors commented to refine it. S.A., D.D., J.S., and J.K. led the FinnTwin12 cohort. R.T., V.M., B.R., and M.F. enriched urban physical exposures and T.P. managed the FinnTwin12 data. Z.W. performed the analysis and wrote the original draft. All authors reviewed the draft and approved for the submission.

## Ethical requirement

The ethics committee of the Department of Public Health of the University of Helsinki (Helsinki, Finland) and the Institutional Review Board of Indiana University (Bloomington, Indiana, USA) approved the FinnTwin12 study protocol from the start of the cohort. The ethical approval of the ethics committee of the Helsinki University Central Hospital District (HUS) is the most recent and covers the most recent data collection (early midlife) (HUS/2226/2021, dated September 22, 2021). All participants and their parents/legal guardians gave informed written consent to participate in the study. The authors assert that all procedures contributing to this work comply with the ethical standards of the relevant national and institutional committees on human experimentation and with the Helsinki Declaration of 1975, as revised in 2008.

## Data and Code Availability

Code for major analyses is available at https://github.com/doge73/city_urban_PA.

