## Supplementary Note 1-2, Figure 1-3 and Table 1-6 for "The urban physical exposome and leisure-time physical activity in early midlife: a FinnTwin12 study"

Zhiyang Wang et al. – Online supplemental material

Table of contents

Supplemental Note 1: List of questions for physical activity

Supplemental Note 2: Description of obtaining, processing, and calculation of urban exposures

Supplemental Figure 1: Cumulative frequency curves of physical activity measures: (1) total leisure-time physical activity (total LTPA) and its sub-domains (2) leisure-time physical activity without commuting activity (LTPA) and (3) commuting activity

Supplemental Figure 2: the matrix of pairwise SHAP interaction values from the XGBoost model with the outcome of total LTPA

Supplemental Figure 3: the matrix of pairwise SHAP interaction values from the XGBoost model with the outcome of LTPA

Supplemental Table 1: Description of urban physical exposures

Supplemental Table 2: Characteristics of demographic and behavior variables among all available participants from the early midlife follow-up

Supplemental Table 3: Results of the sex-stratified linear regression between urban cluster and physical activity measures

Supplemental Table 4: Results of the linear regression between urban physical exposures and physical activity measures. Age and sex were adjusted.

Supplemental Table 5: Importance rank for the XGBoost model of total LTPA (reported test and two extra tests)

Supplemental Table 6: Importance rank for the XGBoost model of LTPA (reported test and two extra tests)

Supplemental Note 1: List of questions for physical activity

How often do you engage in physical activity during your leisure time?^Frequency^

1 Not at all
2 Less than once a month
3 1–2 times a month
4 Once a week
5 2–3 times a week
6 4–5 times a week
7 Nearly every day
8 Several times a day

Is your physical activity during leisure time about as tiring on average as:^Intensity^

1 Walking
2 Alternatively walking and jogging
3 Jogging (light run)
4 Running

How long does one session of physical activity last on average?^Duration^

1 Less than half an hour
2 Half an hour to less than one hour
3 One hour to less than two hours
4 Two hours or more

How much of your daily journey to work/study is spent in walking, cycling, running and/or cross-country skiing?^Commuting activity^

1 Less than 15 minutes
2 15 minutes to less than half an hour
3 Half an hour to less than one hour
4 One hour or more
5 I am presently not at work or studying

Supplemental Note 2: Description of obtaining, processing, and calculation of urban exposures

Data about the greenspaces, parks, and street networks is from OpenStreetMap (OSM)^1^, which is a valuable source of urban morphology and land use data with high granularity. Its well-documented taxonomy also ensures that the physical environment features in the study areas. Greenspaces are interconnected areas tagged as “greenspace”, including parks, but also forests, meadows, grasslands, etc., of 0.5 hectares in size and larger; normal parks are interconnected areas tagged as “parks” in OSM of 0.5 hectares in size and larger; Pocket parks are interconnected areas tagged as “parks” in OSM up to 0.5 hectares in size. Mean, median, minimum, maximum, and summary sizes and counts within an area of a buffer of 300, 500, and 800 meter walking distance are used to characterize the three types of areas. To estimate the street network, the street network is represented as a graph with nodes as intersections and edges as streets. The betweenness centrality score reflects the distance traveled from one street intersection to all others and is calculated by determining the proportion of shortest paths between all node pairs that pass through that edge^2^. The intersection closeness score reflects the distance traveled from one street intersection to all others and is the average of the shortest path between this node and all the other nodes in the graph is the closeness centrality of each node. The sinuosity score reflects the straightness of a street segment by quantifying how much longer it is relative to a straight line between its start and end points. A line’s sinuosity is defined as the ratio of its curvilinear length and its Euclidean distance.

The percentage of area covered by buildings in the three buffer areas and the percentage of impervious area captures the percentage of soil sealing. Impervious areas are characterized by the substitution of the original (semi-) natural land cover or water surface with an artificial, often impervious cover. These indicators are derived from the Impervious Surface Area (ISA)^3^.

The distance, type, and size of the closest green and blue spaces came from the Europe-wide Urban Atlas (UA)^4^ and CORINE Land Cover^5^ were used to extract maps of urban and natural green and blue spaces. We decided to use these two datasets due to the highest resolution of UA and high coverage of CORINE with the scarce resolution. The EU defines the closest green and blue spaces as living within 300 m of a public open area with more than 5000 m^2^ (or within a 15-minute walk).

There are two types of vegetation index: Normalized Difference Vegetation Index (NDVI) and Modified Soil Adjusted Vegetation Index (MSAVI) to characterize the vegetation density within buffer areas of 100, 300, and 500 m radii around the geocodes. NDVI quantifies vegetation greenness and is useful in understanding vegetation density and assessing changes in plant health^6^. It is calculated as a ratio between the red (R) and near-infrared (NIR) values in a traditional fashion^6^. MSAVI minimizes the effect of bare soil on the Soil Adjusted Vegetation Index, which corrects NDVI for the influence of soil brightness in areas where vegetative cover is low^7^. It is calculated as a ratio between the R and NIR values with an inductive function applied to maximize the reduction of soil effects on the vegetation signal^7^. Their source is the U.S. Geological Survey Landsat data via Google Earth Engine.

Land use gives the percentage of all types of land use within an area of a buffer of 100, 300, and 500 meter radius, obtained from UA^4^. The following land use categories were created, by grouping the land use categories available in UA: “high density residential”, “low density residential”, “industrial, commercial, public, military and private units”, “infrastructure”, “urban green”, “agricultural green” ,“natural green”, “water”. The Minimum Mapping Unit is 0.25 ha.

Population density is counted within the three buffer areas around the geocode, derived as the weighted sum of the population count indicated in each pixel where the buffer area intersects the population raster data. Worldpop’s (WP) estimation models have been developed, refined, and implemented to produce global multi-temporal 100x100 m datasets for each year^8^. The assumption is that no settlement dataset is accurate enough to identify all residential settlements/buildings globally. The modeling is unconstrained, making predictions about population numbers for all 100x100 m of land grid cells globally for each year through disaggregating a census database.

Roads are divided into 4 categories: "Major Roads", "Roads", "Streets" and "Any Road", and indicators of distance to the closest road type and total length of roads within buffer areas of 100, 300, and 500 meters radius to characterize proximity and characteristics of surrounding road. The count of road junctions within buffer areas of 100, 300, and 500 meters radius (for any category of road) is used to characterize the connectivity of streets, roads, and major roads. The count of public transport stops within buffer areas of 100, 300, and 500 meters radius (each line and direction are counted separately and the length of public transport lines within the three buffers (each line and direction are counted separately) are used to characterize accessibility of public transport stops and lines. Quality assessment involves two indicators: saturation and growth. The saturation is the ratio between the count of public transport lines at one point in time and the count of public transport lines and roads' cumulative length expected at a saturated level. The higher the saturation indicator, the closer to completeness is the dataset. The growth is the relative variation in the count of public transport lines and roads' cumulative length over one year. A low growth indicator generally indicates that the "creation" phase (addition of missing features) passed and the dataset is now being "maintained". These data is derived from OSM through ohsome platform after quality assessment^9^.

The elevation and slope within three buffer areas around the geocode are derived from the EU-DEM (digital elevation model) with overall high fundamental vertical accuracy^10^. It ensures that water features are adequately represented and consistent with the hydrography layer^10^. In areas above 60 degrees north, the EU-DEM generation process is supported by other DEM data sources provided by the Joint Research Center’s Global Surface Water dataset. Water features are flattened (oceans, lakes) and stepped (rivers) based on the hydrography data^10^.

Tree cover indicators estimate the percentage of horizontal ground in the 3 buffer areas at 30 m resolution covered by woody vegetation greater than 5 meters in height, and they are from the Global Forest Cover Change data products through NASA^11^.

Supplemental Figure 1: Cumulative frequency curves of physical activity measures: (1) total leisure-time physical activity (total LTPA) and its sub-domains (2) leisure-time physical activity without commuting activity (LTPA) and (3) commuting activity

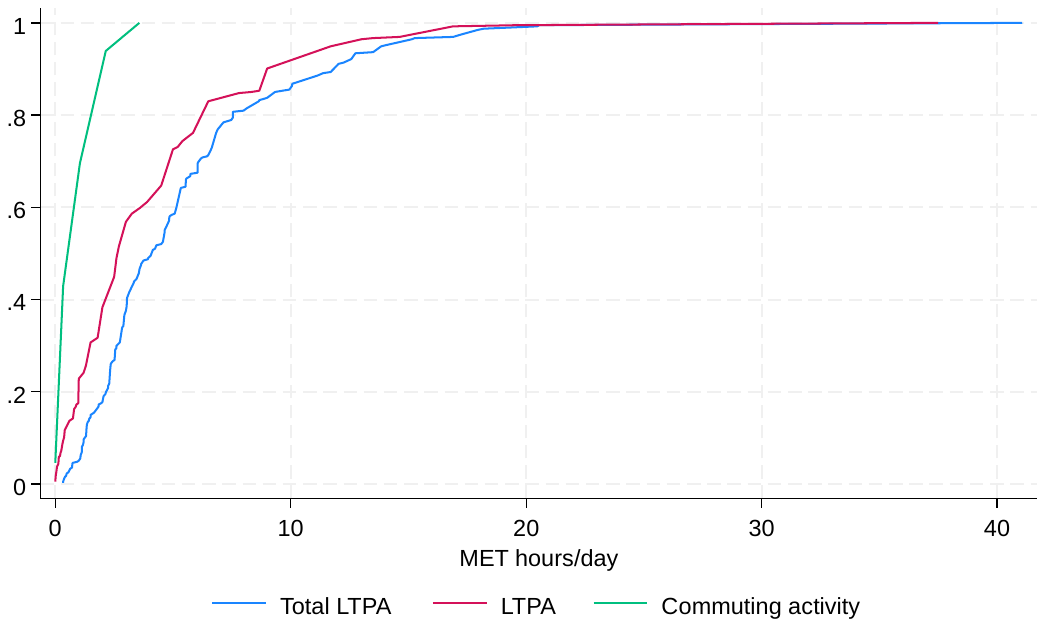

Supplemental Figure 2: the matrix of pairwise SHAP interaction values from the XGBoost model with the outcome of total LTPA

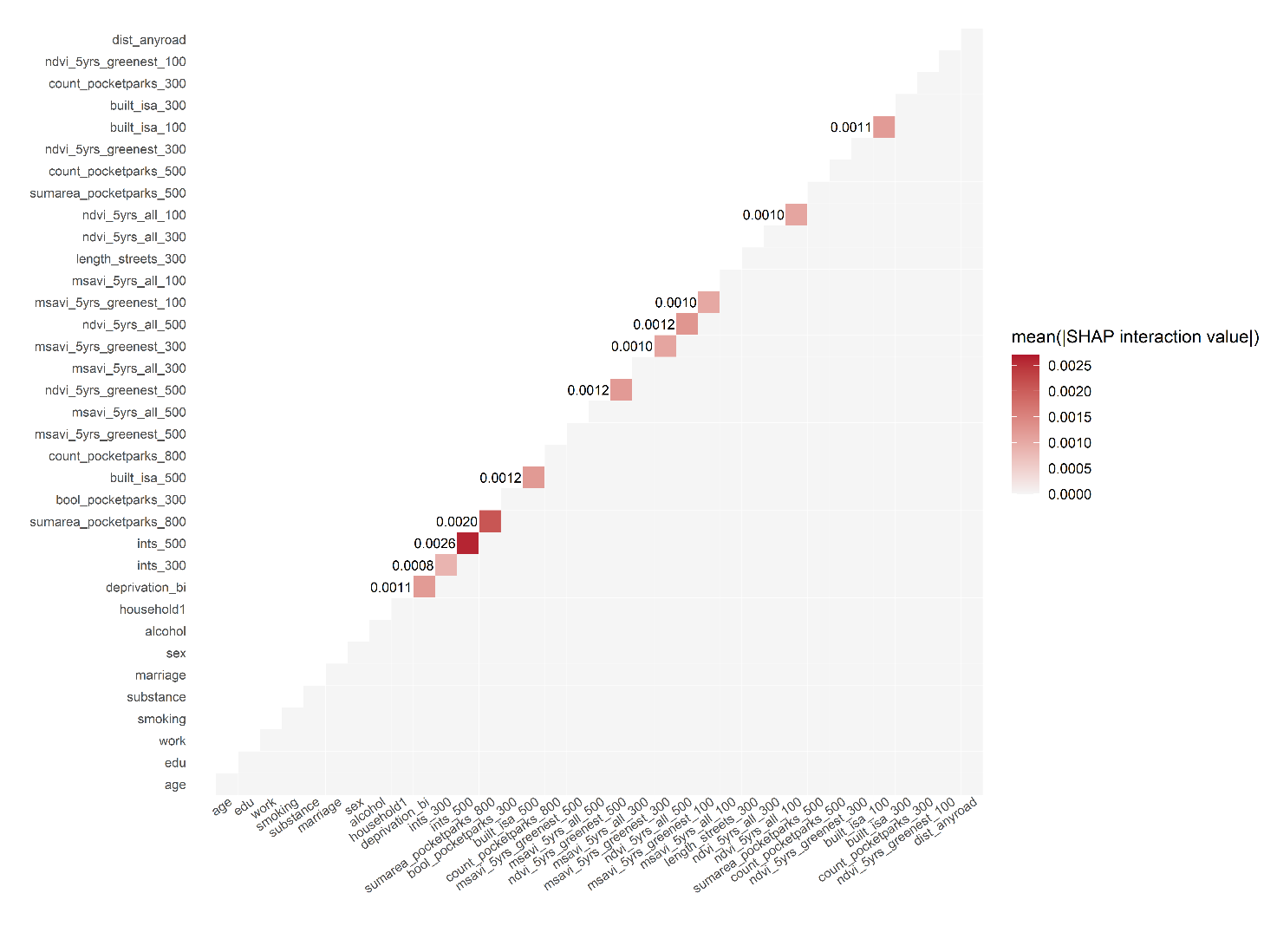

Supplemental Figure 3: the matrix of pairwise SHAP interaction values from the XGBoost model with the outcome of LTPA

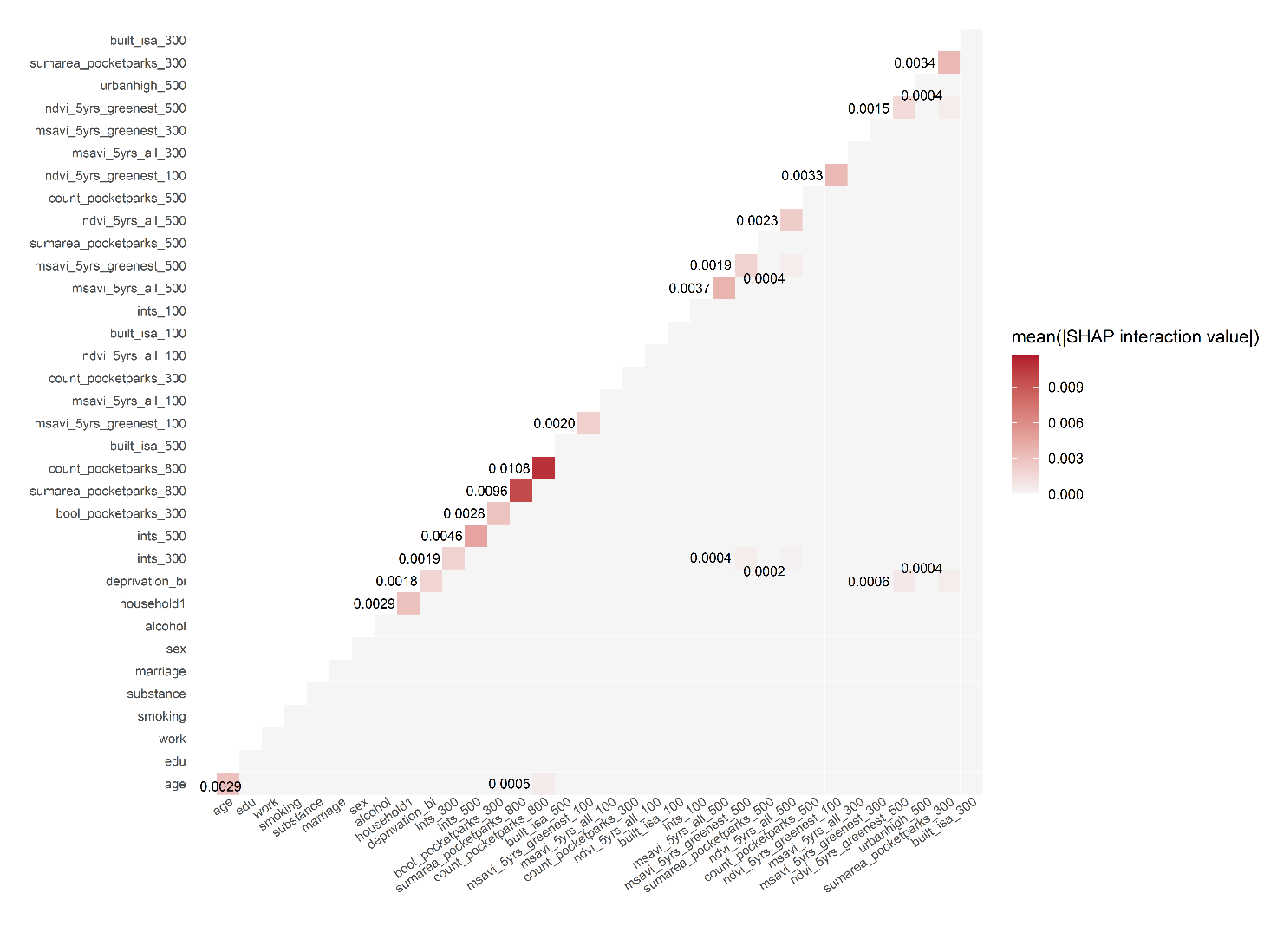

Supplemental Table 1: Description of urban physical exposures

| **Urban exposures** | **Year** | **Unit** | **Mean (SD) / N. (%)** | **Missing N.** | **Description** |
| --- | --- | --- | --- | --- | --- |
| **meanarea_normalparks_300** | 2023 | Square kilometer | 0.28 (1.24) | 0 | The mean area of all interconnected normal parks within a 300 m walking distance from the geo-coordinate, including greenspace areas partly within a 300 m walking distance while further extending beyond walking distance. |
| **medianarea_normalparks_300** | 2023 | Square kilometer | 0.25 (1.20) | 0 | The median area of all interconnected normal parks within a 300 m walking distance from the geo-coordinate, including greenspace areas partly within a 300 m walking distance while further extending beyond walking distance. |
| **minarea_normalparks_300** | 2023 | Square kilometer | 0.18 (1.10) | 0 | The area of the smallest interconnected normal parks within a 300 m walking distance from the geo-coordinate. |
| **maxarea_normalparks_300** | 2023 | Square kilometer | 0.42 (1.78) | 0 | The area of the largest interconnected normal parks within a 300 m walking distance from the geo-coordinate. |
| **sumarea_normalparks_300** | 2023 | Square kilometer | 0.44 (1.78) | 0 | The total area of all interconnected normal parks within a 300 m walking distance from the geo-coordinate, including greenspace areas partly within a 300 m walking distance while further extending beyond walking distance. |
| **count_normalparks_300** | 2023 | / | 1.66 (0.83) | 0 | The count of normal parks within a 300 m walking distance from the geo-coordinate. |
| **sumarea_pocketparks_300** | 2023 | Square kilometer | 0.00 (0.00) | 0 | The total area of all interconnected pocket parks within a 300 m walking distance from the geo-coordinate, including greenspace areas partly within a 300 m walking distance while further extending beyond walking distance. |
| **count_pocketparks_300** | 2023 | / | 0.47 (0.92) | 0 | The count of pocket parks within a 300 m walking distance from the geo-coordinate. |
| **bool_pocketparks_300** | 2023 | / |  | 0 | Whether or not there are pocket parks within a 300 m walking distance from the geo-coordinate. |
| False |  |  | 277 (70.30) |  |  |
| True |  |  | 117 (29.70) |  |  |
| **meanarea_greenspaces_300** | 2023 | Square kilometer | 0.40 (1.33) | 0 | The mean area of all interconnected green spaces within a 300 m walking distance from the geo-coordinate, including greenspace areas partly within a 300 m walking distance while further extending beyond walking distance. |
| **medianarea_greenspaces_300** | 2023 | Square kilometer | 0.28 (1.18) | 0 | The median area of all interconnected green spaces within a 300 m walking distance from the geo-coordinate, including greenspace areas partly within a 300 m walking distance while further extending beyond walking distance. |
| **minarea_greenspaces_300** | 2023 | Square kilometer | 0.14 (0.65) | 0 | The area of the smallest interconnected green spaces within a 300 m walking distance from the geo-coordinate. |
| **maxarea_greenspaces_300** | 2023 | Square kilometer | 0.82 (2.89) | 0 | The area of the largest interconnected green spaces within a 300 m walking distance from the geo-coordinate. |
| **sumarea_greenspaces_300** | 2023 | Square kilometer | 0.86 (2.91) | 0 | The total area of all interconnected green spaces within a 300 m walking distance from the geo-coordinate, including greenspace areas partly within a 300 m walking distance while further extending beyond walking distance. |
| **count_greenspaces_300** | 2023 | / | 2.13 (1.04) | 0 | The count of green spacess within a 300 m walking distance from the geo-coordinate. |
| **meanarea_normalparks_500** | 2023 | Square kilometer | 0.25 (1.03) | 0 | The mean area of all interconnected normal parks within a 500 m walking distance from the geo-coordinate, including greenspace areas partly within a 500 m walking distance while further extending beyond walking distance. |
| **medianarea_normalparks_500** | 2023 | Square kilometer | 0.15 (0.91) | 0 | The median area of all interconnected normal parks within a 500 m walking distance from the geo-coordinate, including greenspace areas partly within a 500 m walking distance while further extending beyond walking distance. |
| **minarea_normalparks_500** | 2023 | Square kilometer | 0.11 (0.89) | 0 | The area of the smallest interconnected normal parks within a 500 m walking distance from the geo-coordinate. |
| **maxarea_normalparks_500** | 2023 | Square kilometer | 0.54 (2.01) | 0 | The area of the largest interconnected normal parks within a 500 m walking distance from the geo-coordinate. |
| **sumarea_normalparks_500** | 2023 | Square kilometer | 0.59 (2.02) | 0 | The total area of all interconnected normal parks within a 500 m walking distance from the geo-coordinate, including greenspace areas partly within a 500 m walking distance while further extending beyond walking distance. |
| **count_normalparks_500** | 2023 | / | 3.02 (1.67) | 0 | The count of normal parks within a 500 m walking distance from the geo-coordinate. |
| **sumarea_pocketparks_500** | 2023 | Square kilometer | 0.00 (0.00) | 0 | The total area of all interconnected pocket parks within a 500 m walking distance from the geo-coordinate, including greenspace areas partly within a 500 m walking distance while further extending beyond walking distance. |
| **count_pocketparks_500** | 2023 | / | 1.09 (1.61) | 0 | The count of pocket parks within a 500 m walking distance from the geo-coordinate. |
| **bool_pocketparks_500** | 2023 | / |  | 0 | Whether or not there are pocket parks within a 500 m walking distance from the geo-coordinate. |
| False |  |  | 193 (48.98) |  |  |
| True |  |  | 201 (51.02) |  |  |
| **meanarea_greenspaces_500** | 2023 | Square kilometer | 0.52 (2.12) | 0 | The mean area of all interconnected green spaces within a 500 m walking distance from the geo-coordinate, including greenspace areas partly within a 500 m walking distance while further extending beyond walking distance. |
| **medianarea_greenspaces_500** | 2023 | Square kilometer | 0.18 (1.02) | 0 | The median area of all interconnected green spaces within a 500 m walking distance from the geo-coordinate, including greenspace areas partly within a 500 m walking distance while further extending beyond walking distance. |
| **minarea_greenspaces_500** | 2023 | Square kilometer | 0.09 (0.55) | 0 | The area of the smallest interconnected green spaces within a 500 m walking distance from the geo-coordinate. |
| **maxarea_greenspaces_500** | 2023 | Square kilometer | 1.66 (6.77) | 0 | The area of the largest interconnected green spaces within a 500 m walking distance from the geo-coordinate. |
| **sumarea_greenspaces_500** | 2023 | Square kilometer | 1.78 (6.79) | 0 | The total area of all interconnected green spaces within a 500 m walking distance from the geo-coordinate, including greenspace areas partly within a 500 m walking distance while further extending beyond walking distance. |
| **count_greenspaces_500** | 2023 | / | 4.06 (1.72) | 0 | The count of green spaces within a 500 m walking distance from the geo-coordinate. |
| **meanarea_normalparks_800** | 2023 | Square kilometer | 0.24 (0.65) | 0 | The mean area of all interconnected normal parks within an 800 m walking distance from the geo-coordinate, including greenspace areas partly within an 800 m walking distance while further extending beyond walking distance. |
| **medianarea_normalparks_800** | 2023 | Square kilometer | 0.06 (0.29) | 0 | The median area of all interconnected normal parks within an 800 m walking distance from the geo-coordinate, including greenspace areas partly within an 800 m walking distance while further extending beyond walking distance. |
| **minarea_normalparks_800** | 2023 | Square kilometer | 0.02 (0.08) | 0 | The area of the smallest interconnected normal parks within an 800 m walking distance from the geo-coordinate. |
| **maxarea_normalparks_800** | 2023 | Square kilometer | 0.95 (2.74) | 0 | The area of the largest interconnected normal parks within an 800 m walking distance from the geo-coordinate. |
| **sumarea_normalparks_800** | 2023 | Square kilometer | 1.07 (2.77) | 0 | The total area of all interconnected normal parks within an 800 m walking distance from the geo-coordinate, including greenspace areas partly within an 800 m walking distance while further extending beyond walking distance. |
| **count_normalparks_800** | 2023 | / | 5.57 (3.10) | 0 | The count of normal parks within an 800 m walking distance from the geo-coordinate. |
| **sumarea_pocketparks_800** | 2023 | Square kilometer | 0.01 (0.01) | 0 | The total area of all interconnected pocket parks within an 800 m walking distance from the geo-coordinate, including greenspace areas partly within an 800 m walking distance while further extending beyond walking distance. |
| **count_pocketparks_800** | 2023 | / | 2.30 (2.68) | 0 | The count of pocket parks within an 800 m walking distance from the geo-coordinate. |
| **bool_pocketparks_800** | 2023 | / |  | 0 | Whether or not there are pocket parks within an 800 m walking distance from the geo-coordinate. |
| False |  |  | 125 (31.73) |  |  |
| True |  |  | 269 (68.27) |  |  |
| **meanarea_greenspaces_800** | 2023 | Square kilometer | 0.46 (1.87) | 0 | The mean area of all interconnected green spaces within an 800 m walking distance from the geo-coordinate, including greenspace areas partly within an 800 m walking distance while further extending beyond walking distance. |
| **medianarea_greenspaces_800** | 2023 | Square kilometer | 0.10 (0.63) | 0 | The median area of all interconnected green spaces within an 800 m walking distance from the geo-coordinate, including greenspace areas partly within an 800 m walking distance while further extending beyond walking distance. |
| **minarea_greenspaces_800** | 2023 | Square kilometer | 0.01 (0.03) | 0 | The area of the smallest interconnected green spaces within an 800 m walking distance from the geo-coordinate. |
| **maxarea_greenspaces_800** | 2023 | Square kilometer | 2.22 (7.00) | 0 | The area of the largest interconnected green spaces within an 800 m walking distance from the geo-coordinate. |
| **sumarea_greenspaces_800** | 2023 | Square kilometer | 2.57 (7.37) | 0 | The total area of all interconnected green spaces within an 800 m walking distance from the geo-coordinate, including greenspace areas partly within an 800 m walking distance while further extending beyond walking distance. |
| **count_greenspaces_800** | 2023 | / | 7.86 (2.96) | 0 | The count of green spaces within an 800 m walking distance from the geo-coordinate. |
| **wavg_street_betweenness_100** | 2023 | / | 12600000 (20100000) | 0 | The average street betweenness scores of every street segment within a 100 m from each geo-coordinate weighted according to the street segment's length. |
| **wavg_street_sinuosity_100** | 2023 | / | 1.09 (0.14) | 0 | The average street sinuosity scores of every street segment within a 100 m from each geo-coordinate weighted according to the street segment’s length. |
| **mean_intersection_closeness_100** | 2023 | / | 0.00 (0.02) | 0 | The mean street intersection closeness scores of every street segment within a 100 m from each geo-coordinate. |
| **wavg_street_betweenness_300** | 2023 | / | 15300000 (16100000) | 0 | The average street betweenness scores of every street segment within a 300 m from each geo-coordinate weighted according to the street segment's length. |
| **wavg_street_sinuosity_300** | 2023 | / | 1.08 (0.06) | 0 | The average street sinuosity scores of every street segment within a 300 m from each geo-coordinate weighted according to the street segment’s length. |
| **mean_intersection_closeness_300** | 2023 | / | 0.00 (0.01) | 0 | The mean street intersection closeness scores of every street segment within a 300 m from each geo-coordinate. |
| **nearest_street_length** | 2023 | Meter | 68.85 (53.45) | 0 | The length of the nearest street segment to each geo-coordinate. |
| **nearest_street_betweenness** | 2023 | / | 3317406 (10200000) | 0 | The betweenness score of the nearest street segment to each geo-coordinate. |
| **nearest_street_sinuosity** | 2023 | / | 1.12 (0.53) | 2 | The sinuosity score of the nearest street segment to each geo-coordinate. |
| **nearest_street_dist** | 2023 | Meter | 12.58 (6.11) | 0 | The distance between each geo-coordinate and the nearest street segment. |
| **nearest_street_highway_footway** | 2023 | / |  | 0 | The type of the nearest street segment to each geo-coordinate is footway. |
| no |  |  | 183 (46.45) |  |  |
| yes |  |  | 211 (53.55) |  |  |
| **nearest_street_highway_service** | 2023 | / |  | 0 | The type of the nearest street segment to each geo-coordinate is service way. |
| no |  |  | 302 (76.65) |  |  |
| yes |  |  | 92 (23.35) |  |  |
| **nearest_street_highway_other** | 2023 | / |  | 0 | The type of the nearest street segment to each geo-coordinate is other types. |
| no |  |  | 296 (75.13) |  |  |
| yes |  |  | 98 (24.87) |  |  |
| **built_isa_100** | 2018 | / | 53.78 (22.15) | 0 | Percentage (%) of impervious area within a buffer area of 100 m radius around each geo-coordinate (source: High Resolution Layer Imperviousness). |
| **built_isa_300** | 2018 | / | 48.20 (20.19) | 0 | Percentage (%) of impervious area within a buffer area of 300 m radius around each geo-coordinate (source: High Resolution Layer Imperviousness). |
| **built_isa_500** | 2018 | / | 45.00 (19.17) | 0 | Percentage (%) of impervious area within a buffer area of 500 m radius around each geo-coordinate (source: High Resolution Layer Imperviousness). |
| **dist_green_ua** | 2018 | Meter | 0.09 (0.07) | 8 | Euclidean distance to the closest green space (source: Urban Atlas). Land use categories for green: (‘14100’, ‘14200’, ‘21000’, ‘22000’, ‘23000’, ‘24000’,‘25000’, ‘31000’, ‘32000’, ‘40000’) |
| **item_green_ua** | 2018 | / |  | 0 | Land use category of the closest green space (source: Urban Atlas). |
| Other |  |  | 148 (37.56) |  |  |
| Green urban areas |  |  | 246 (62.44) |  |  |
| **dist_blue_ua** | 2018 | Meter | 0.93 (0.77) | 8 | Euclidean distance to the closest blue space (source: Urban Atlas). Land use categories for blue: (‘50000’). |
| **size_blue_ua** | 2018 | Square meter | 10.22 (29.55) | 8 | Area of the closest blue space (source: Urban Atlas). |
| **dist_major_green_ua** | 2018 | Meter | 0.09 (0.07) | 8 | Euclidean distance to the closest major green space (source: Urban Atlas). Land use categories for green: (‘14100’, ‘14200’, ‘21000’, ‘22000’, ‘23000’, ‘24000’,‘25000’, ‘31000’, ‘32000’, ‘40000’). Major means >5000 m2. |
| **item_major_green_ua** | 2018 | / |  | 0 | Land use category of the closest major green space (source: Urban Atlas). Major means >5000 m2 |
| Other |  |  | 153 (38.83) |  |  |
| Green urban areas |  |  | 241 (61.17) |  |  |
| **dist_major_blue_ua** | 2018 | Meter | 0.93 (0.77) | 8 | Euclidean distance to the closest major green space (source: Urban Atlas). Land use categories for green: (‘14100’, ‘14200’, ‘21000’, ‘22000’, ‘23000’, ‘24000’,‘25000’, ‘31000’, ‘32000’, ‘40000’). Major means >5000 m2. |
| **size_major_blue_ua** | 2018 | Square meter | 10.22 (29.55) | 8 | Euclidean distance to the closest blue space (source: Urban Atlas). Land use categories for blue: (‘50000’). Major means >5000 m2. |
| **dist_green_clc** | 2018 | Meter | 0.59 (0.48) | 0 | Euclidean distance to the closest green space (source: Corine Land Cover). Land use categories for green: (‘141’, ‘142’, ‘211’, ‘212’, ‘213’, ‘221’, ‘222’, ‘223’, ‘231’, ‘241’, ‘242’, ‘243’, ‘244’, ‘311’, ‘312’, ‘313’, ‘321’, ‘322’, ‘323’, ‘324’, ‘333’, ‘334’, ‘335’, ‘411’, ‘412’). |
| **dist_blue_clc** | 2018 | Meter | 1.23 (1.67) | 0 | Euclidean distance to the closest blue space (source: Corine Land Cover). Land use categories for blue: (‘331’, ‘421’, ‘422’, ‘423’, ‘511’, ‘512’, ‘521’, ‘522’, ‘523’). |
| **size_blue_clc** | 2018 | Square meter | 30794.35 (27030.29) | 0 | Area of the closest blue space (source: Corine Land Cover). |
| **item_blue_clc** | 2018 | / |  | 0 | Land use category of the closest blue (source: Corine Land Cover). |
| Water bodies and courses |  |  | 118 (29.95) |  |  |
| Sea and ocean |  |  | 276 (70.05) |  |  |
| **access_green_ua_300** | 2018 | / |  | 8 | Access to a major green space, ie. the presence of a major blue space within 300 m from geocode (Euclidean distance) (source: Urban Atlas). |
| No |  |  | 4 (1.04) |  |  |
| Yes |  |  | 382 (98.96) |  |  |
| **access_blue_ua_300** | 2018 | / |  | 8 | Access to a major blue space, ie. the presence of a major blue space within 300 m from geocode (Euclidean distance) (source: Urban Atlas). |
| No |  |  | 311 (80.57) |  |  |
| Yes |  |  | 75 (19.43) |  |  |
| **urbanhigh_100** | 2018 | / | 0.46 (0.32) | 8 | Percentage of high-density residential land use type within a 100 m around each geo-coordinate. |
| **urbanlow_100** | 2018 | / | 0.19 (0.29) | 8 | Percentage of low-density residential land use type within a 100 m around each geo-coordinate. |
| **com_ind_100** | 2018 | / | 0.10 (0.19) | 8 | Percentage of commercial and industrial land use type within a 100 m around each geo-coordinate. |
| **infrast_100** | 2018 | / | 0.12 (0.12) | 8 | Percentage of infrastructure land use type within a 100 m around each geo-coordinate. |
| **green_urb_100** | 2018 | / | 0.08 (0.12) | 8 | Percentage of urban green land use type within a 100 m around each geo-coordinate. |
| **agric_100** | 2018 | / | 0.00 (0.03) | 8 | Percentage of agricultural land use type within a 100 m around each geo-coordinate. |
| **natural_100** | 2018 | / | 0.04 (0.10) | 8 | Percentage of natural land use type within a 100 m around each geo-coordinate. |
| **water_100** | 2018 | / | 0.00 (0.03) | 8 | Percentage of water land use type within a 100 m around each geo-coordinate. |
| **urbanhigh_300** | 2018 | / | 0.33 (0.20) | 8 | Percentage of high-density residential land use type within a 300 m around each geo-coordinate. |
| **urbanlow_300** | 2018 | / | 0.15 (0.19) | 8 | Percentage of low-density residential land use type within a 300 m around each geo-coordinate. |
| **com_ind_300** | 2018 | / | 0.15 (0.13) | 8 | Percentage of commercial and industrial land use type within a 300 m around each geo-coordinate. |
| **infrast_300** | 2018 | / | 0.13 (0.09) | 8 | Percentage of infrastructure land use type within a 300 m around each geo-coordinate. |
| **green_urb_300** | 2018 | / | 0.13 (0.11) | 8 | Percentage of urban green land use type within a 300 m around each geo-coordinate. |
| **agric_300** | 2018 | / | 0.01 (0.04) | 8 | Percentage of agricultural land use type within a 300 m around each geo-coordinate. |
| **natural_300** | 2018 | / | 0.07 (0.13) | 8 | Percentage of natural land use type within a 300 m around each geo-coordinate. |
| **water_300** | 2018 | / | 0.02 (0.07) | 8 | Percentage of water land use type within a 300 m around each geo-coordinate. |
| **urbanhigh_500** | 2018 | / | 0.27 (0.16) | 8 | Percentage of high-density residential land use type within a 500 m around each geo-coordinate. |
| **urbanlow_500** | 2018 | / | 0.14 (0.16) | 8 | Percentage of low-density residential land use type within a 500 m around each geo-coordinate. |
| **com_ind_500** | 2018 | / | 0.15 (0.11) | 8 | Percentage of commercial and industrial land use type within a 500 m around each geo-coordinate. |
| **infrast_500** | 2018 | / | 0.13 (0.08) | 8 | Percentage of infrastructure land use type within a 500 m around each geo-coordinate. |
| **green_urb_500** | 2018 | / | 0.15 (0.10) | 8 | Percentage of urban green land use type within a 500 m around each geo-coordinate. |
| **agric_500** | 2018 | / | 0.01 (0.04) | 8 | Percentage of agricultural land use type within a 500 m around each geo-coordinate. |
| **natural_500** | 2018 | / | 0.09 (0.14) | 8 | Percentage of natural land use type within a 500 m around each geo-coordinate. |
| **water_500** | 2018 | / | 0.04 (0.09) | 8 | Percentage of water land use type within a 500 m around each geo-coordinate. |
| **pop_wp_100** | 2018 | / | 98.21 (43.11) | 0 | Population count within a 100 m buffer around each geo-coordinate (source: WorldPop). |
| **pop_wp_300** | 2018 | / | 859.80 (382.09) | 0 | Population count within a 300 m buffer around each geo-coordinate (source: WorldPop). |
| **pop_wp_500** | 2018 | / | 2321.24 (1055.13) | 0 | Population count within a 500 m buffer around each geo-coordinate (source: WorldPop). |
| **dist_streets** | 2018 | Meter | 0.04 (0.03) | 0 | Euclidian distance to the closest road of category 'streets'. |
| **length_streets_100** | 2018 | Meter | 0.28 (0.19) | 0 | Total length of road sections of category 'streets' within a 100 m radius buffer. |
| **length_streets_300** | 2018 | Meter | 2.05 (1.04) | 0 | Total length of road sections of category 'streets' within a 300 m radius buffer. |
| **length_streets_500** | 2018 | Meter | 5.11 (2.42) | 0 | Total length of road sections of category 'streets' within a 500 m radius buffer. |
| **dist_roads** | 2018 | Meter | 0.15 (0.15) | 0 | Euclidian distance to the closest road of category 'roads'. |
| **length_roads_100** | 2018 | Meter | 0.10 (0.14) | 0 | Total length of road sections of category 'roads' within a 100 m radius buffer. |
| **length_roads_300** | 2018 | Meter | 0.90 (0.61) | 0 | Total length of road sections of category 'roads' within a 300 m radius buffer. |
| **length_roads_500** | 2018 | Meter | 2.30 (1.28) | 0 | Total length of road sections of category 'roads' within a 500 m radius buffer. |
| **dist_majorroads** | 2018 | Meter | 0.65 (1.46) | 0 | Euclidian distance to the closest road of category 'major_roads'. |
| **length_majorroads_100** | 2018 | Meter | 0.03 (0.09) | 0 | Total length of road sections of category 'major_roads' within a 100 m radius buffer. |
| **length_majorroads_300** | 2018 | Meter | 0.47 (0.74) | 0 | Total length of road sections of category 'major_roads' within a 300 m radius buffer. |
| **length_majorroads_500** | 2018 | Meter | 1.49 (1.68) | 0 | Total length of road sections of category 'major_roads' within a 500 m radius buffer. |
| **dist_anyroad** | 2018 | Meter | 0.03 (0.03) | 0 | Euclidian distance to the closest road of category 'any_road'. |
| **ne_dem_100** | 2018 | Meter | 42.10 (35.18) | 0 | Elevation within a 100 m buffer around each geo-coordinate. |
| **ne_dem_300** | 2018 | Meter | 41.55 (35.24) | 0 | Elevation within a 300 m buffer around each geo-coordinate. |
| **ne_dem_500** | 2018 | Meter | 41.24 (35.16) | 0 | Elevation within a 500 m buffer around each geo-coordinate. |
| **ne_slo_100** | 2018 | / | 2.02 (2.93) | 0 | Slope within a 100 m buffer around each geo-coordinate. |
| **ne_slo_300** | 2018 | / | 2.02 (2.02) | 0 | Slope within a 300 m buffer around each geo-coordinate. |
| **ne_slo_500** | 2018 | / | 2.14 (1.68) | 0 | Slope within a 500 m buffer around each geo-coordinate. |
| **msavi_5yrs_all_100** | 2018 | / | 0.15 (0.07) | 0 | 5-year moving average of modified soil adjusted vegetation index (MSAVI) within a 100 m buffer around each geo-coordinate using all available images during the whole year. |
| **msavi_5yrs_greenest_100** | 2018 | / | 0.18 (0.08) | 0 | 5-year moving average of MSAVI during the greenest season (May, June, July, and August) within a 100 m buffer around each geo-coordinate using all available images during the whole year. |
| **msavi_5yrs_all_300** | 2018 | / | 0.17 (0.06) | 0 | 5-year moving average of MSAVI within a 300 m buffer around each geo-coordinate using all available images during the whole year. |
| **msavi_5yrs_greenest_300** | 2018 | / | 0.20 (0.07) | 0 | 5-year moving average of MSAVI during the greenest season (May, June, July, and August) within a 300 m buffer around each geo-coordinate using all available images during the whole year. |
| **msavi_5yrs_all_500** | 2018 | / | 0.17 (0.06) | 0 | 5-year moving average of MSAVI within a 500 m buffer around each geo-coordinate using all available images during the whole year. |
| **msavi_5yrs_greenest_500** | 2018 | / | 0.21 (0.07) | 0 | 5-year moving average of MSAVI during the greenest season (May, June, July, and August) within a 500 m buffer around each geo-coordinate using all available images during the whole year. |
| **ndvi_5yrs_all_100** | 2018 | / | 0.34 (0.13) | 0 | 5-year moving average of normalized difference vegetation index (NDVI) within a 100 m buffer around each geo-coordinate using all available images during the whole year. |
| **ndvi_5yrs_greenest_100** | 2018 | / | 0.39 (0.14) | 0 | 5-year moving average of NDVI during the greenest season (May, June, July, and August) within a 100 m buffer around each geo-coordinate using all available images during the whole year. |
| **ndvi_5yrs_all_300** | 2018 | / | 0.37 (0.12) | 0 | 5-year moving average of NDVI within a 300 m buffer around each geo-coordinate using all available images during the whole year. |
| **ndvi_5yrs_greenest_300** | 2018 | / | 0.41 (0.13) | 0 | 5-year moving average of NDVI during the greenest season (May, June, July, and August) within a 300 m buffer around each geo-coordinate using all available images during the whole year. |
| **ndvi_5yrs_all_500** | 2018 | / | 0.38 (0.12) | 0 | 5-year moving average of NDVI within a 500 m buffer around each geo-coordinate using all available images during the whole year. |
| **ndvi_5yrs_greenest_500** | 2018 | / | 0.43 (0.13) | 0 | 5-year moving average of NDVI during the greenest season (May, June, July, and August) within a 500 m buffer around each geo-coordinate using all available images during the whole year. |
| **ints_100** | 2018 | / | 2.45 (2.91) | 0 | Count of road junctions of any type within a 100 m buffer around each geo-coordinate. |
| **ints_300** | 2018 | / | 19.26 (13.16) | 0 | Count of road junctions of any type within a 300 m buffer around each geo-coordinate. |
| **ints_500** | 2018 | / | 48.94 (29.16) | 0 | Count of road junctions of any type within a 500 m buffer around each geo-coordinate. |
| **stops_100** | 2018 | / | 2.71 (8.64) | 0 | Count of public transport stops within a 100 m buffer around each geo-coordinate. |
| **lines_100** | 2018 | Meter | 1426.34 (2815.6) | 0 | Total length of public transport lines within a 100 m radius buffer. |
| **stops_300** | 2018 | / | 20.12 (26.11) | 0 | Count of public transport stops within a 300 m buffer around each geo-coordinate. |
| **lines_300** | 2018 | Meter | 14178.21 (16862.66) | 0 | Total length of public transport lines within a 300 m radius buffer. |
| **stops_500** | 2018 | / | 50.00 (56.66) | 0 | Count of public transport stops within a 500 m buffer around each geo-coordinate. |
| **lines_500** | 2018 | Meter | 37182.43 (37234.88) | 0 | Total length of public transport lines within a 500 m radius buffer. |
| **treecover_100** | 2015 | / | 22.33 (7.63) | 4 | Percentage of area covered by trees within a 100 m buffer around each geo-coordinate. |
| **treecover_300** | 2015 | / | 23.68 (7.65) | 4 | Percentage of area covered by trees within a 300 m buffer around each geo-coordinate. |
| **treecover_500** | 2015 | / | 24.08 (7.65) | 4 | Percentage of area covered by trees within a 500 m buffer around each geo-coordinate. |

Supplemental Table 2: Characteristics of sociodemographic and behavior variables among included and excluded participants

| **Characteristics** | **Included participants** | | **Exclude participants** | | **P-value*** |
| --- | --- | --- | --- | --- | --- |
|  | N. | N. (%) / Mean (SD) | N. | N. (%) / Mean (SD) |  |
| **Sex** | 394 |  | 1726 |  | 0.06 |
| Male |  | 179 (45.4) |  | 696 (40.3) |  |
| Female |  | 215 (54.6) |  | 1030 (59.7) |  |
| **Work** | 394 |  | 1710 |  | 0.46 |
| Not working or other situation |  | 51 (12.9) |  | 246 (14.4) |  |
| Currently work |  | 343 (87.1) |  | 1464 (85.6) |  |
| **Education** | 394 |  | 1710 |  | <0.001 |
| Post-secondary or lower |  | 84 (21.3) |  | 676 (39.5) |  |
| Bachelor/equivalent or above |  | 310 (78.7) |  | 1034 (60.5) |  |
| **Marital status** | 394 |  | 1680 |  | 0.08 |
| No |  | 97 (24.6) |  | 345 (20.5) |  |
| Married, steady relationship, or living together |  | 297 (75.4) |  | 1335 (79.5) |  |
| **Age (years)** | 394 | 37.1 (1.5) | 1726 | 37.2 (1.5) | 0.60 |
| **Illicit substance use** | 394 |  | 1673 |  | <0.001 |
| Never |  | 204 (51.8) |  | 1114 (66.6) |  |
| At least once |  | 190 (48.2) |  | 559 (33.4) |  |
| **Ever smoker (smoked over 100 cigarettes lifetime)** | 394 |  | 1671 |  | 0.28 |
| No |  | 216 (54.8) |  | 866 (51.8) |  |
| Yes |  | 178 (45.2) |  | 805 (48.2) |  |
| **Alcohol** | 394 |  | 1677 |  | 0.01 |
| Monthly or less, or even never |  | 164 (41.6) |  | 824 (49.1) |  |
| 2-4 times a month or more |  | 230 (58.4) |  | 853 (50.9) |  |

* P-values were calculated by analysis of variance or chi-square test.

Supplemental Table 3: Results of the sex-stratified linear regression between urban cluster and physical activity measures

| **Outcome  (log-transformed)** | **Characteristics** | **Beta (95% CI)** | | |
| --- | --- | --- | --- | --- |
|  |  | Model 1 ^a^ | Model 2 ^b^ | Model 3 ^c^ |
| *Male (n=178)* | | | | |
| **Total LTPA** | **Urban cluster** |  |  |  |
|  | 1 (original city center) | Ref. | Ref. | Ref. |
|  | 2 (new city center) | -0.06 (-0.16, 0.04) | -0.06 (-0.17, 0.04) | -0.06 (-0.18, 0.05) |
|  | 3 (suburban) | -0.16 (-0.27, -0.06)* | -0.16 (-0.27, -0.05)* | -0.16 (-0.29, -0.03)* |
| **LTPA** | **Urban cluster** |  |  |  |
|  | 1 (original city center) | Ref. | Ref. | Ref. |
|  | 2 (new city center) | -0.04 (-0.17, 0.08) | -0.04 (-0.17, 0.09) | -0.04 (-0.19, 0.10) |
|  | 3 (suburban) | -0.17 (-0.300, -0.03)* | -0.16 (-0.30, -0.02)* | -0.17 (-0.33, 0.00) |
| **Commuting activity** | **Urban cluster** |  |  |  |
|  | 1 (original city center) | Ref. | Ref. | Ref. |
|  | 2 (new city center) | -0.04 (-0.19, 0.10) | -0.05 (-0.12, 0.02) | -0.04 (-0.12, 0.03) |
|  | 3 (suburban) | -0.17 (-0.33, 0.00) | -0.07 (-0.14, 0.01) | -0.06 (-0.15, 0.02) |
| *Female (n=213)* | | | | |
| **Total LTPA** | **Urban cluster** |  |  |  |
|  | 1 (original city center) | Ref. | Ref. | Ref. |
|  | 2 (new city center) | -0.06 (-0.16, 0.03) | -0.04 (-0.14, 0.05) | -0.04 (-0.14, 0.07) |
|  | 3 (suburban) | -0.14 (-0.24, -0.03)* | -0.11 (-0.22, 0.01) | -0.10 (-0.24, 0.04) |
| **LTPA** | **Urban cluster** |  |  |  |
|  | 1 (original city center) | Ref. | Ref. | Ref. |
|  | 2 (new city center) | -0.08 (-0.19, 0.03) | -0.07 (-0.18, 0.04) | -0.07 (-0.20, 0.05) |
|  | 3 (suburban) | -0.18 (-0.30, -0.06)* | -0.16 (-0.28, -0.03) | -0.16 (-0.32, 0.00) |
| **Commuting activity** | **Urban cluster** |  |  |  |
|  | 1 (original city center) | Ref. | Ref. | Ref. |
|  | 2 (new city center) | -0.00 (-0.06, 0.06) | 0.01 (-0.05, 0.07) | 0.03 (-0.03, 0.09) |
|  | 3 (suburban) | -0.02 (-0.09, 0.04) | 0.00 (-0.06, 0.07) | 0.03 (-0.05, 0.10) |

* P<0.05

^a^ Adjusted for age, sex, education, work, and marital status

^b^ Based on model 1, additionally adjusted for smoking, alcohol drinking, and illicit substance use

^c^ Based on model 2, additionally adjusted for neighborhood deprivation level and the proportion of single households in the neighborhood

Supplemental Table 4: Results of the linear regression between urban physical exposures and physical activity measures. Age and sex were adjusted.

| **Urban exposure** | **Total LTPA** | | **LTPA** | | **Commuting activity** | |
| --- | --- | --- | --- | --- | --- | --- |
|  | Beta (95% CI) | P-value | Beta (95% CI) | P-value | Beta (95% CI) | P-value |
| **meanarea_normalparks_300** | -0.0000 (-0.0000, 0.0000) | 0.8643 | 0.0000 (-0.0000, 0.0000) | 0.7394 | -0.0000 (-0.0000, 0.0000) | 0.2369 |
| **medianarea_normalparks_300** | 0.0000 (-0.0000, 0.0000) | 0.8280 | 0.0000 (-0.0000, 0.0000) | 0.5065 | -0.0000 (-0.0000, 0.0000) | 0.3629 |
| **minarea_normalparks_300** | 0.0000 (-0.0000, 0.0000) | 0.9605 | 0.0000 (-0.0000, 0.0000) | 0.7168 | -0.0000 (-0.0000, 0.0000) | 0.6230 |
| **maxarea_normalparks_300** | -0.0000 (-0.0000, 0.0000) | 0.5935 | 0.0000 (-0.0000, 0.0000) | 0.9775 | -0.0000 (-0.0000, 0.0000) | 0.1174 |
| **sumarea_normalparks_300** | -0.0043 (-0.0196, 0.0111) | 0.5880 | 0.0002 (-0.0179, 0.0183) | 0.9814 | -0.0081 (-0.0181, 0.0020) | 0.1173 |
| **count_normalparks_300** | -0.0036 (-0.0368, 0.0296) | 0.8320 | -0.0034 (-0.0424, 0.0355) | 0.8629 | 0.0069 (-0.0149, 0.0287) | 0.5348 |
| **sumarea_pocketparks_300** | 12.7723 (2.1989, 23.3457) | 0.0184 | 16.4579 (4.0529, 28.8629) | 0.0097 | 0.2259 (-6.7610, 7.2128) | 0.9495 |
| **count_pocketparks_300** | 0.0413 (0.0119, 0.0707) | 0.0062 | 0.0516 (0.0171, 0.0861) | 0.0036 | 0.0043 (-0.0152, 0.0238) | 0.6643 |
| **bool_pocketparks_300** |  |  |  |  |  |  |
| False | Ref. | | Ref. | | Ref. | |
| True | 0.1000 (0.0409, 0.1592) | 0.0010 | 0.1330 (0.0638, 0.2022) | 0.0002 | -0.0096 (-0.0489, 0.0298) | 0.6331 |
| **meanarea_greenspaces_300** | -0.0108 (-0.0313, 0.0097) | 0.3013 | -0.0069 (-0.0310, 0.0172) | 0.5755 | -0.0104 (-0.0238, 0.0030) | 0.1295 |
| **medianarea_greenspaces_300** | -0.0125 (-0.0357, 0.0106) | 0.2904 | -0.0097 (-0.0370, 0.0175) | 0.4840 | -0.0073 (-0.0225, 0.0079) | 0.3455 |
| **minarea_greenspaces_300** | -0.0013 (-0.0436, 0.0409) | 0.9501 | 0.0025 (-0.0471, 0.0521) | 0.9199 | -0.0018 (-0.0295, 0.0259) | 0.9001 |
| **maxarea_greenspaces_300** | -0.0043 (-0.0137, 0.0052) | 0.3782 | -0.0022 (-0.0133, 0.0089) | 0.6991 | -0.0057 (-0.0118, 0.0005) | 0.0738 |
| **sumarea_greenspaces_300** | -0.0042 (-0.0136, 0.0052) | 0.3816 | -0.0021 (-0.0132, 0.0089) | 0.7026 | -0.0056 (-0.0117, 0.0005) | 0.0746 |
| **count_greenspaces_300** | -0.0009 (-0.0272, 0.0253) | 0.9446 | 0.0002 (-0.0307, 0.0310) | 0.9924 | 0.0036 (-0.0136, 0.0208) | 0.6822 |
| **meanarea_normalparks_500** | -0.0112 (-0.0377, 0.0152) | 0.4064 | -0.0045 (-0.0356, 0.0266) | 0.7756 | -0.0135 (-0.0308, 0.0038) | 0.1268 |
| **medianarea_normalparks_500** | -0.0080 (-0.0381, 0.0221) | 0.6035 | -0.0018 (-0.0373, 0.0336) | 0.9194 | -0.0119 (-0.0316, 0.0078) | 0.2382 |
| **minarea_normalparks_500** | -0.0040 (-0.0349, 0.0268) | 0.7977 | 0.0022 (-0.0340, 0.0384) | 0.9061 | -0.0103 (-0.0305, 0.0099) | 0.3200 |
| **maxarea_normalparks_500** | -0.0091 (-0.0227, 0.0045) | 0.1885 | -0.0063 (-0.0223, 0.0097) | 0.4428 | -0.0073 (-0.0162, 0.0016) | 0.1092 |
| **sumarea_normalparks_500** | -0.0094 (-0.0229, 0.0041) | 0.1742 | -0.0066 (-0.0225, 0.0093) | 0.4169 | -0.0072 (-0.0160, 0.0017) | 0.1121 |
| **count_normalparks_500** | -0.0050 (-0.0214, 0.0114) | 0.5509 | -0.0062 (-0.0254, 0.0130) | 0.5286 | 0.0039 (-0.0068, 0.0146) | 0.4774 |
| **sumarea_pocketparks_500** | 8.7316 (2.7173, 14.7458) | 0.0047 | 10.107 (3.0385, 17.1755) | 0.0053 | 2.8776 (-1.0989, 6.8541) | 0.1569 |
| **count_pocketparks_500** | 0.0243 (0.0075, 0.0411) | 0.0048 | 0.0281 (0.0083, 0.0478) | 0.0056 | 0.0059 (-0.0052, 0.0171) | 0.2963 |
| **bool_pocketparks_500** |  |  |  |  |  |  |
| False | Ref. | | Ref. | | Ref. | |
| True | 0.0647 (0.0104, 0.1190) | 0.0200 | 0.0740 (0.0102, 0.1378) | 0.0236 | 0.0205 (-0.0153, 0.0563) | 0.2634 |
| **meanarea_greenspaces_500** | 0.0020 (-0.0109, 0.0149) | 0.7620 | 0.0035 (-0.0117, 0.0186) | 0.6513 | -0.0004 (-0.0089, 0.0080) | 0.9230 |
| **medianarea_greenspaces_500** | -0.0027 (-0.0295, 0.0240) | 0.8412 | 0.0019 (-0.0295, 0.0334) | 0.9041 | -0.0066 (-0.0242, 0.0109) | 0.4594 |
| **minarea_greenspaces_500** | 0.0089 (-0.0409, 0.0587) | 0.7268 | 0.0116 (-0.0469, 0.0701) | 0.6975 | 0.0075 (-0.0252, 0.0401) | 0.6548 |
| **maxarea_greenspaces_500** | -0.0002 (-0.0042, 0.0039) | 0.9321 | 0.0000 (-0.0047, 0.0048) | 0.9859 | -0.0003 (-0.0030, 0.0023) | 0.8028 |
| **sumarea_greenspaces_500** | -0.0002 (-0.0042, 0.0038) | 0.9136 | -0.0000 (-0.0048, 0.0047) | 0.9922 | -0.0003 (-0.0030, 0.0023) | 0.8145 |
| **count_greenspaces_500** | -0.0089 (-0.0248, 0.0070) | 0.2734 | -0.0084 (-0.0271, 0.0103) | 0.3787 | -0.0021 (-0.0125, 0.0083) | 0.6934 |
| **meanarea_normalparks_800** | 0.0001 (-0.0420, 0.0423) | 0.9956 | 0.0159 (-0.0336, 0.0654) | 0.5291 | -0.0214 (-0.0490, 0.0062) | 0.1290 |
| **medianarea_normalparks_800** | -0.0189 (-0.1139, 0.0761) | 0.6972 | -0.0052 (-0.1168, 0.1064) | 0.9274 | -0.0229 (-0.0852, 0.0394) | 0.4722 |
| **minarea_normalparks_800** | 0.1046 (-0.2400, 0.4492) | 0.5521 | 0.1578 (-0.2470, 0.5626) | 0.4452 | -0.1507 (-0.3764, 0.0751) | 0.1916 |
| **maxarea_normalparks_800** | 0.0033 (-0.0067, 0.0133) | 0.5149 | 0.0070 (-0.0047, 0.0188) | 0.2404 | -0.0034 (-0.0100, 0.0031) | 0.3069 |
| **sumarea_normalparks_800** | 0.0031 (-0.0067, 0.0130) | 0.5336 | 0.0068 (-0.0048, 0.0184) | 0.2498 | -0.0034 (-0.0099, 0.0031) | 0.3072 |
| **count_normalparks_800** | 0.0049 (-0.0039, 0.0138) | 0.2755 | 0.0063 (-0.0041, 0.0167) | 0.2376 | 0.0021 (-0.0037, 0.0079) | 0.4853 |
| **sumarea_pocketparks_800** | 6.1224 (2.6278, 9.6170) | 0.0007 | 7.4937 (3.3931, 11.5943) | 0.0004 | 0.9352 (-1.3902, 3.2606) | 0.4310 |
| **count_pocketparks_800** | 0.0169 (0.0069, 0.0270) | 0.0011 | 0.0203 (0.0085, 0.0322) | 0.0008 | 0.0024 (-0.0043, 0.0091) | 0.4751 |
| **bool_pocketparks_800** |  |  |  |  |  |  |
| False | Ref. | | Ref. | | Ref. | |
| True | 0.0500 (-0.0085, 0.1084) | 0.0948 | 0.0545 (-0.0143, 0.1232) | 0.1212 | 0.0138 (-0.0247, 0.0523) | 0.4821 |
| **meanarea_greenspaces_800** | 0.0072 (-0.0074, 0.0218) | 0.3370 | 0.0097 (-0.0074, 0.0269) | 0.2679 | 0.0009 (-0.0087, 0.0105) | 0.8581 |
| **medianarea_greenspaces_800** | 0.0205 (-0.0232, 0.0642) | 0.3592 | 0.0274 (-0.0240, 0.0787) | 0.2968 | -0.0024 (-0.0311, 0.0263) | 0.8711 |
| **minarea_greenspaces_800** | 0.3974 (-0.5198, 1.3145) | 0.3963 | 0.4927 (-0.5847, 1.5702) | 0.3706 | -0.4772 (-1.0776, 0.1232) | 0.1201 |
| **maxarea_greenspaces_800** | 0.0005 (-0.0034, 0.0045) | 0.7847 | 0.0011 (-0.0035, 0.0057) | 0.6454 | -0.0002 (-0.0027, 0.0024) | 0.9068 |
| **sumarea_greenspaces_800** | 0.0008 (-0.0029, 0.0045) | 0.6861 | 0.0013 (-0.0031, 0.0056) | 0.5614 | -0.0001 (-0.0025, 0.0023) | 0.9330 |
| **count_greenspaces_800** | -0.0041 (-0.0133, 0.0052) | 0.3864 | -0.0032 (-0.0141, 0.0076) | 0.5608 | -0.0016 (-0.0076, 0.0045) | 0.6116 |
| **wavg_street_betweenness_100** | 0.0000 (0.0000, 0.0000) | 0.0451 | 0.0000 (0.0000, 0.0000) | 0.0144 | -0.0000 (-0.0000, 0.0000) | 0.9885 |
| **wavg_street_sinuosity_100** | 0.0889 (-0.1070, 0.2848) | 0.3742 | 0.1371 (-0.0929, 0.3671) | 0.2434 | -0.0602 (-0.1887, 0.0684) | 0.3594 |
| **mean_intersection_closeness_100** | -0.4628 (-1.8986, 0.9731) | 0.5279 | -0.2132 (-1.901, 1.4745) | 0.8045 | -0.4612 (-1.4026, 0.4802) | 0.3376 |
| **wavg_street_betweenness_300** | 0.0000 (-0.0000, 0.0000) | 0.0826 | 0.0000 (-0.0000, 0.0000) | 0.0641 | 0.0000 (-0.0000, 0.0000) | 0.3533 |
| **wavg_street_sinuosity_300** | -0.2429 (-0.7102, 0.2243) | 0.3088 | -0.2785 (-0.8276, 0.2705) | 0.3207 | -0.0598 (-0.3668, 0.2471) | 0.7027 |
| **mean_intersection_closeness_300** | 1.3665 (-3.0292, 5.7622) | 0.5427 | 1.8330 (-3.3309, 6.9970) | 0.4870 | 1.0685 (-1.8150, 3.9520) | 0.4681 |
| **nearest_street_length** | 0.0002 (-0.0003, 0.0007) | 0.4524 | 0.0002 (-0.0004, 0.0008) | 0.4306 | -0.0002 (-0.0005, 0.0001) | 0.2678 |
| **nearest_street_betweenness** | 0.0000 (-0.0000, 0.0000) | 0.3982 | 0.0000 (-0.0000, 0.0000) | 0.3892 | -0.0000 (-0.0000, 0.0000) | 0.9106 |
| **nearest_street_sinuosity** | 0.0059 (-0.0454, 0.0572) | 0.8211 | -0.0065 (-0.0668, 0.0539) | 0.8340 | 0.0162 (-0.0174, 0.0498) | 0.3455 |
| **nearest_street_dist** | -0.0035 (-0.0080, 0.0010) | 0.1304 | -0.0032 (-0.0085, 0.0021) | 0.2402 | -0.0033 (-0.0063, -0.0004) | 0.0267 |
| **nearest_street_highway_footway** |  |  |  |  |  |  |
| no | Ref. | | Ref. | | Ref. | |
| yes | 0.0438 (-0.0108, 0.0983) | 0.1167 | 0.0422 (-0.0220, 0.1063) | 0.1987 | 0.0358 (0.0001, 0.0715) | 0.0501 |
| **nearest_street_highway_service** |  |  |  |  |  |  |
| no | Ref. | | Ref. | | Ref. | |
| yes | -0.0399 (-0.1052, 0.0254) | 0.2320 | -0.0417 (-0.1185, 0.0351) | 0.2877 | -0.0243 (-0.0672, 0.0185) | 0.2668 |
| **nearest_street_highway_other** |  |  |  |  |  |  |
| no | Ref. | | Ref. | | Ref. | |
| yes | -0.0174 (-0.0815, 0.0467) | 0.5947 | -0.0160 (-0.0913, 0.0594) | 0.6782 | -0.0237 (-0.0657, 0.0183) | 0.2688 |
| **built_isa_100** | 0.0017 (0.0005, 0.0030) | 0.0054 | 0.0021 (0.0007, 0.0035) | 0.0042 | 0.0003 (-0.0005, 0.0011) | 0.4912 |
| **built_isa_300** | 0.0019 (0.0006, 0.0033) | 0.0058 | 0.0021 (0.0005, 0.0037) | 0.0097 | 0.0006 (-0.0002, 0.0015) | 0.1565 |
| **built_isa_500** | 0.0024 (0.0010, 0.0038) | 0.0010 | 0.0026 (0.0010, 0.0043) | 0.0020 | 0.0009 (0.0000, 0.0018) | 0.0620 |
| **dist_green_ua** | -0.3487 (-0.7645, 0.0671) | 0.1011 | -0.5632 (-1.0503, -0.0761) | 0.0240 | 0.1211 (-0.1524, 0.3946) | 0.3860 |
| **item_green_ua** |  |  |  |  |  |  |
| Other | Ref. | | Ref. | | Ref. | |
| Green urban areas | 0.0431 (-0.0132, 0.0993) | 0.1342 | 0.0518 (-0.0143, 0.1178) | 0.1253 | 0.0137 (-0.0232, 0.0507) | 0.4669 |
| **dist_blue_ua** | 0.0062 (-0.0298, 0.0423) | 0.7356 | 0.0056 (-0.0368, 0.0479) | 0.7966 | 0.0056 (-0.0180, 0.0293) | 0.6415 |
| **size_blue_ua** | 0.0008 (-0.0002, 0.0017) | 0.1098 | 0.0010 (-0.0001, 0.0021) | 0.0736 | 0.0000 (-0.0006, 0.0007) | 0.9090 |
| **dist_major_green_ua** | -0.2127 (-0.6089, 0.1836) | 0.2935 | -0.4222 (-0.8865, 0.0421) | 0.0755 | 0.1580 (-0.1018, 0.4179) | 0.2341 |
| **item_major_green_ua** |  |  |  |  |  |  |
| Other | Ref. | | Ref. | | Ref. | |
| Green urban areas | 0.0421 (-0.0138, 0.0980) | 0.1408 | 0.0538 (-0.0119, 0.1194) | 0.1092 | 0.0138 (-0.0230, 0.0506) | 0.4625 |
| **dist_major_blue_ua** | 0.0072 (-0.0289, 0.0433) | 0.6966 | 0.0067 (-0.0357, 0.0491) | 0.7565 | 0.0058 (-0.0179, 0.0295) | 0.6312 |
| **size_major_blue_ua** | 0.0008 (-0.0002, 0.0017) | 0.1099 | 0.0010 (-0.0001, 0.0021) | 0.0736 | 0.0000 (-0.0006, 0.0007) | 0.9092 |
| **dist_green_clc** | 0.0384 (-0.0184, 0.0951) | 0.1856 | 0.0403 (-0.0264, 0.1070) | 0.2371 | 0.0145 (-0.0228, 0.0518) | 0.4473 |
| **dist_blue_clc** | -0.0042 (-0.0206, 0.0122) | 0.6150 | -0.0032 (-0.0225, 0.0161) | 0.7439 | 0.0002 (-0.0106, 0.0110) | 0.9722 |
| **size_blue_clc** | 0.0000 (-0.0000, 0.0000) | 0.2366 | 0.0000 (-0.0000, 0.0000) | 0.1439 | -0.0000 (-0.0000, 0.0000) | 0.7564 |
| **item_blue_clc** |  |  |  |  |  |  |
| Water bodies and courses | Ref. | | Ref. | | Ref. | |
| Sea and ocean | 0.0491 (-0.0106, 0.1088) | 0.1076 | 0.0624 (-0.0077, 0.1325) | 0.0816 | 0.0130 (-0.0263, 0.0522) | 0.5183 |
| **access_green_ua_300** |  |  |  |  |  |  |
| No | Ref. | | Ref. | | Ref. | |
| Yes | 0.0930 (-0.1794, 0.3653) | 0.5038 | 0.1975 (-0.122, 0.5171) | 0.2264 | -0.0606 (-0.2393, 0.1181) | 0.5068 |
| **access_blue_ua_300** |  |  |  |  |  |  |
| No | Ref. | | Ref. | | Ref. | |
| Yes | -0.0229 (-0.0934, 0.0476) | 0.5240 | -0.0428 (-0.1255, 0.0400) | 0.3119 | 0.0105 (-0.0357, 0.0568) | 0.6560 |
| **urbanhigh_100** | 0.0366 (-0.0503, 0.1235) | 0.4093 | 0.0423 (-0.0598, 0.1444) | 0.4171 | 0.0009 (-0.0562, 0.0579) | 0.9766 |
| **urbanlow_100** | -0.0884 (-0.1829, 0.0060) | 0.0673 | -0.1109 (-0.2218, 0.0000) | 0.0508 | -0.0164 (-0.0787, 0.0458) | 0.6047 |
| **com_ind_100** | 0.0756 (-0.0718, 0.2229) | 0.3154 | 0.1056 (-0.0675, 0.2786) | 0.2325 | 0.0098 (-0.0870, 0.1066) | 0.8431 |
| **infrast_100** | 0.1286 (-0.1036, 0.3608) | 0.2783 | 0.0960 (-0.1770, 0.3691) | 0.4910 | 0.1014 (-0.0508, 0.2537) | 0.1924 |
| **green_urb_100** | 0.1183 (-0.1125, 0.3492) | 0.3157 | 0.1399 (-0.1314, 0.4111) | 0.3129 | 0.0375 (-0.1142, 0.1891) | 0.6286 |
| **agric_100** | -0.9858 (-1.8648, -0.1068) | 0.0285 | -1.1304 (-2.1634, -0.0973) | 0.0326 | -0.2075 (-0.7874, 0.3724) | 0.4835 |
| **natural_100** | -0.1312 (-0.3940, 0.1316) | 0.3286 | -0.0910 (-0.4000, 0.2180) | 0.5642 | -0.0678 (-0.2403, 0.1047) | 0.4416 |
| **water_100** | 0.2437 (-0.6632, 1.1506) | 0.5987 | 0.3433 (-0.7220, 1.4087) | 0.5280 | -0.0160 (-0.6112, 0.5793) | 0.9581 |
| **urbanhigh_300** | 0.1557 (0.0188, 0.2926) | 0.0264 | 0.1859 (0.0251, 0.3467) | 0.0240 | 0.0293 (-0.0611, 0.1196) | 0.5258 |
| **urbanlow_300** | -0.1540 (-0.2948, -0.0132) | 0.0327 | -0.1998 (-0.3650, -0.0346) | 0.0183 | -0.0382 (-0.1310, 0.0547) | 0.4207 |
| **com_ind_300** | 0.1252 (-0.0805, 0.3310) | 0.2337 | 0.1418 (-0.1000, 0.3835) | 0.2512 | 0.0567 (-0.0784, 0.1918) | 0.4113 |
| **infrast_300** | 0.1773 (-0.1214, 0.4759) | 0.2453 | 0.1360 (-0.2152, 0.4873) | 0.4483 | 0.1710 (-0.0245, 0.3666) | 0.0873 |
| **green_urb_300** | -0.0961 (-0.3535, 0.1613) | 0.4646 | -0.0385 (-0.3411, 0.2641) | 0.8031 | -0.0697 (-0.2385, 0.0992) | 0.4192 |
| **agric_300** | -0.3659 (-1.0958, 0.3639) | 0.3264 | -0.6071 (-1.4636, 0.2493) | 0.1655 | 0.1817 (-0.2974, 0.6608) | 0.4578 |
| **natural_300** | -0.1407 (-0.3547, 0.0733) | 0.1983 | -0.1078 (-0.3596, 0.1440) | 0.4019 | -0.1091 (-0.2494, 0.0312) | 0.1283 |
| **water_300** | -0.0008 (-0.3736, 0.3721) | 0.9969 | -0.0636 (-0.5016, 0.3744) | 0.7762 | 0.0607 (-0.1838, 0.3053) | 0.6268 |
| **urbanhigh_500** | 0.2152 (0.0462, 0.3843) | 0.0130 | 0.2645 (0.0660, 0.4630) | 0.0093 | 0.0473 (-0.0644, 0.1590) | 0.4072 |
| **urbanlow_500** | -0.1983 (-0.3666, -0.0300) | 0.0215 | -0.2474 (-0.4450, -0.0499) | 0.0145 | -0.0609 (-0.1719, 0.0502) | 0.2833 |
| **com_ind_500** | 0.2242 (-0.0262, 0.4745) | 0.0800 | 0.2698 (-0.0243, 0.5639) | 0.0729 | 0.0824 (-0.0823, 0.2471) | 0.3274 |
| **infrast_500** | 0.3009 (-0.0346, 0.6365) | 0.0795 | 0.2319 (-0.1632, 0.627) | 0.2507 | 0.2702 (0.0509, 0.4896) | 0.0162 |
| **green_urb_500** | -0.1048 (-0.3770, 0.1675) | 0.4511 | -0.0731 (-0.3931, 0.2469) | 0.6546 | -0.0406 (-0.2193, 0.1381) | 0.6561 |
| **agric_500** | -0.2036 (-0.9399, 0.5327) | 0.5881 | -0.3826 (-1.2472, 0.482) | 0.3863 | 0.1781 (-0.3049, 0.6610) | 0.4703 |
| **natural_500** | -0.1326 (-0.3292, 0.0640) | 0.1871 | -0.1154 (-0.3466, 0.1159) | 0.3287 | -0.1129 (-0.2417, 0.0159) | 0.0865 |
| **water_500** | -0.0902 (-0.3841, 0.2038) | 0.5481 | -0.1624 (-0.5076, 0.1828) | 0.3570 | 0.0417 (-0.1512, 0.2346) | 0.6719 |
| **pop_wp_100** | 0.0005 (-0.0002, 0.0011) | 0.1486 | 0.0006 (-0.0002, 0.0013) | 0.1353 | 0.0002 (-0.0002, 0.0006) | 0.4127 |
| **pop_wp_300** | 0.0001 (-0.0000, 0.0001) | 0.1402 | 0.0001 (-0.0000, 0.0002) | 0.1105 | 0.0000 (-0.0000, 0.0001) | 0.5065 |
| **pop_wp_500** | 0.0000 (-0.0000, 0.0000) | 0.1019 | 0.0000 (-0.0000, 0.0001) | 0.0651 | 0.0000 (-0.0000, 0.0000) | 0.5958 |
| **dist_streets** | -0.8124 (-1.6296, 0.0048) | 0.0521 | -0.8067 (-1.7683, 0.1548) | 0.1009 | -0.4052 (-0.9425, 0.1321) | 0.1402 |
| **length_streets_100** | 0.1441 (-0.0029, 0.2910) | 0.0554 | 0.1362 (-0.0368, 0.3091) | 0.1236 | 0.0727 (-0.0239, 0.1693) | 0.1409 |
| **length_streets_300** | 0.0403 (0.0142, 0.0664) | 0.0027 | 0.0398 (0.0090, 0.0706) | 0.0117 | 0.0203 (0.0031, 0.0375) | 0.0214 |
| **length_streets_500** | 0.0147 (0.0035, 0.0260) | 0.0103 | 0.0161 (0.0030, 0.0293) | 0.0168 | 0.0050 (-0.0024, 0.0124) | 0.1839 |
| **dist_roads** | -0.1831 (-0.3689, 0.0028) | 0.0543 | -0.2495 (-0.4675, -0.0315) | 0.0255 | 0.0474 (-0.075, 0.1699) | 0.4483 |
| **length_roads_100** | 0.1874 (-0.0036, 0.3784) | 0.0552 | 0.2861 (0.0624, 0.5097) | 0.0126 | -0.0972 (-0.2227, 0.0283) | 0.1299 |
| **length_roads_300** | 0.0329 (-0.0117, 0.0775) | 0.1485 | 0.0476 (-0.0047, 0.0999) | 0.0754 | -0.0025 (-0.0319, 0.0268) | 0.8657 |
| **length_roads_500** | 0.0227 (0.0013, 0.0441) | 0.0380 | 0.0257 (0.0005, 0.0508) | 0.0461 | 0.0104 (-0.0037, 0.0244) | 0.1502 |
| **dist_majorroads** | -0.0021 (-0.0209, 0.0168) | 0.8306 | 0.0010 (-0.0211, 0.0231) | 0.9298 | -0.0066 (-0.0189, 0.0058) | 0.2986 |
| **length_majorroads_100** | 0.2563 (-0.0410, 0.5536) | 0.0919 | 0.3281 (-0.0210, 0.6772) | 0.0662 | 0.0436 (-0.1521, 0.2393) | 0.6626 |
| **length_majorroads_300** | 0.0227 (-0.0142, 0.0595) | 0.2291 | 0.0357 (-0.0076, 0.0790) | 0.1067 | -0.009 (-0.0332, 0.0152) | 0.4679 |
| **length_majorroads_500** | 0.0166 (0.0004, 0.0328) | 0.0449 | 0.0200 (0.0010, 0.0391) | 0.0395 | 0.002 (-0.0087, 0.0126) | 0.7167 |
| **dist_anyroad** | -1.4154 (-2.4707, -0.3602) | 0.0089 | -1.5598 (-2.8010, -0.3186) | 0.0142 | -0.5138 (-1.2104, 0.1829) | 0.1491 |
| **ne_dem_100** | -0.0003 (-0.0011, 0.0004) | 0.3862 | -0.0006 (-0.0015, 0.0004) | 0.2278 | -0.0000 (-0.0006, 0.0005) | 0.8814 |
| **ne_dem_300** | -0.0003 (-0.0011, 0.0005) | 0.4356 | -0.0005 (-0.0014, 0.0004) | 0.2598 | -0.0000 (-0.0006, 0.0005) | 0.8946 |
| **ne_dem_500** | -0.0003 (-0.0011, 0.0005) | 0.4184 | -0.0005 (-0.0015, 0.0004) | 0.2475 | -0.0000 (-0.0006, 0.0005) | 0.8677 |
| **ne_slo_100** | -0.0012 (-0.0105, 0.0082) | 0.8030 | -0.0020 (-0.0130, 0.0089) | 0.7149 | 0.0007 (-0.0055, 0.0068) | 0.8299 |
| **ne_slo_300** | -0.0006 (-0.0141, 0.0129) | 0.9269 | -0.0017 (-0.0176, 0.0141) | 0.8301 | 0.0017 (-0.0071, 0.0106) | 0.7007 |
| **ne_slo_500** | 0.0037 (-0.0125, 0.0200) | 0.6523 | 0.0054 (-0.0137, 0.0245) | 0.5805 | -0.0007 (-0.0114, 0.0100) | 0.9002 |
| **msavi_5yrs_all_100** | -0.6278 (-1.0341, -0.2216) | 0.0026 | -0.7347 (-1.2120, -0.2574) | 0.0027 | -0.1632 (-0.4324, 0.1059) | 0.2353 |
| **msavi_5yrs_greenest_100** | -0.5452 (-0.8969, -0.1935) | 0.0025 | -0.6422 (-1.0554, -0.2290) | 0.0025 | -0.1302 (-0.3633, 0.1029) | 0.2744 |
| **msavi_5yrs_all_300** | -0.6902 (-1.1277, -0.2526) | 0.0021 | -0.7276 (-1.2429, -0.2123) | 0.0059 | -0.2887 (-0.5778, 0.0005) | 0.0511 |
| **msavi_5yrs_greenest_300** | -0.5936 (-0.9701, -0.2171) | 0.0021 | -0.6192 (-1.0627, -0.1757) | 0.0065 | -0.2481 (-0.4970, 0.0007) | 0.0513 |
| **msavi_5yrs_all_500** | -0.7344 (-1.1872, -0.2816) | 0.0016 | -0.7719 (-1.3053, -0.2384) | 0.0048 | -0.3632 (-0.6619, -0.0644) | 0.0177 |
| **msavi_5yrs_greenest_500** | -0.6402 (-1.0300, -0.2505) | 0.0014 | -0.6615 (-1.1208, -0.2021) | 0.0050 | -0.3199 (-0.5771, -0.0628) | 0.0152 |
| **ndvi_5yrs_all_100** | -0.2949 (-0.4977, -0.0922) | 0.0046 | -0.3511 (-0.5892, -0.1129) | 0.0041 | -0.0652 (-0.1995, 0.0690) | 0.3416 |
| **ndvi_5yrs_greenest_100** | -0.2679 (-0.4592, -0.0767) | 0.0063 | -0.3187 (-0.5434, -0.0941) | 0.0057 | -0.0588 (-0.1854, 0.0677) | 0.3627 |
| **ndvi_5yrs_all_300** | -0.3215 (-0.5394, -0.1036) | 0.0040 | -0.3361 (-0.5927, -0.0795) | 0.0106 | -0.1377 (-0.2816, 0.0061) | 0.0613 |
| **ndvi_5yrs_greenest_300** | -0.2945 (-0.5001, -0.0888) | 0.0053 | -0.3015 (-0.5437, -0.0593) | 0.0152 | -0.1302 (-0.2659, 0.0055) | 0.0608 |
| **ndvi_5yrs_all_500** | -0.3442 (-0.5635, -0.1249) | 0.0022 | -0.3689 (-0.6271, -0.1107) | 0.0054 | -0.1585 (-0.3033, -0.0137) | 0.0325 |
| **ndvi_5yrs_greenest_500** | -0.333 (-0.5436, -0.1223) | 0.0021 | -0.3453 (-0.5935, -0.0972) | 0.0067 | -0.1618 (-0.3007, -0.0228) | 0.0231 |
| **ints_100** | 0.0123 (0.0030, 0.02160) | 0.0100 | 0.0160 (0.0050, 0.0269) | 0.0044 | -0.0013 (-0.0075, 0.0048) | 0.6703 |
| **ints_300** | 0.0043 (0.0023, 0.0064) | 0.0000 | 0.0050 (0.0026, 0.0074) | 0.0000 | 0.0010 (-0.0003, 0.0024) | 0.1429 |
| **ints_500** | 0.0019 (0.0009, 0.0028) | 0.0001 | 0.0021 (0.0010, 0.0032) | 0.0002 | 0.0005 (-0.0001, 0.0011) | 0.0904 |
| **stops_100** | 0.0035 (0.0003, 0.0066) | 0.0309 | 0.0046 (0.0009, 0.0082) | 0.0161 | -0.0004 (-0.0025, 0.0017) | 0.7063 |
| **lines_100** | 0.0000 (-0.0000, 0.0000) | 0.0852 | 0.0000 (0.0000, 0.0000) | 0.0389 | -0.0000 (-0.0000, 0.0000) | 0.8049 |
| **stops_300** | 0.0010 (-0.0001, 0.0020) | 0.0684 | 0.0010 (-0.0002, 0.0023) | 0.1007 | 0.0005 (-0.0002, 0.0012) | 0.1619 |
| **lines_300** | 0.0000 (0.0000, 0.0000) | 0.0471 | 0.0000 (0.0000, 0.0000) | 0.0342 | 0.0000 (-0.0000, 0.0000) | 0.4549 |
| **stops_500** | 0.0005 (0.0000, 0.0010) | 0.0393 | 0.0006 (-0.0000, 0.0011) | 0.0574 | 0.0002 (-0.0001, 0.0006) | 0.1459 |
| **lines_500** | 0.0000 (0.0000, 0.0000) | 0.0250 | 0.0000 (0.0000, 0.0000) | 0.0301 | 0.0000 (-0.0000, 0.0000) | 0.1287 |
| **treecover_100** | -0.0014 (-0.0050, 0.0022) | 0.4608 | -0.0019 (-0.0062, 0.0023) | 0.3705 | -0.0002 (-0.0025, 0.0022) | 0.8847 |
| **treecover_300** | -0.0029 (-0.0064, 0.0007) | 0.1200 | -0.0032 (-0.0074, 0.0010) | 0.1344 | -0.0009 (-0.0032, 0.0015) | 0.4775 |
| **treecover_500** | -0.0034 (-0.0070, 0.0002) | 0.0624 | -0.0037 (-0.0079, 0.0005) | 0.0873 | -0.0013 (-0.0037, 0.0011) | 0.2833 |

Supplemental Table 5: Importance rank for the XGBoost model of total LTPA (reported test and two extra tests)

| Importance rank | Total LTPA | | | | | |
| --- | --- | --- | --- | --- | --- | --- |
|  | Reported test | | Extra test 1 | | Extra test 2 | |
|  | Variables | mean(\|SHAP value\|) | Variables | mean(\|SHAP value\|) | Variables | mean(\|SHAP value\|) |
| 1 | ints_500 | 0.0026 | ints_500 | 0.0068 | ints_300 | 0.0032 |
| 2 | sumarea_pocketparks_800 | 0.0021 | ints_300 | 0.0040 | ints_500 | 0.0031 |
| 3 | ndvi_5yrs_all_500 | 0.0013 | length_streets_300 | 0.0034 | msavi_5yrs_greenest_500 | 0.0024 |
| 4 | built_isa_500 | 0.0012 | sumarea_pocketparks_800 | 0.0030 | ndvi_5yrs_all_100 | 0.0012 |
| 5 | ndvi_5yrs_greenest_500 | 0.0012 | msavi_5yrs_all_500 | 0.0016 | length_streets_300 | 0.0012 |
| 6 | deprivation_bi | 0.0012 | dist_anyroad | 0.0013 | / | / |
| 7 | built_isa_100 | 0.0012 | built_isa_100 | 0.0013 | / | / |
| 8 | msavi_5yrs_greenest_300 | 0.0011 | built_isa_500 | 0.0012 | / | / |
| 9 | ndvi_5yrs_all_100 | 0.0011 | ndvi_5yrs_greenest_300 | 0.0009 | / | / |
| 10 | msavi_5yrs_greenest_100 | 0.0010 | msavi_5yrs_all_300 | 0.0008 | / | / |

Supplemental Table 6: Importance rank for the XGBoost model of LTPA (reported test and two extra tests)

| Importance rank | LTPA | | | | | |
| --- | --- | --- | --- | --- | --- | --- |
|  | Reported test | | Extra test 1 | | Extra test 2 | |
|  | Variables | mean(\|SHAP value\|) | Variables | mean(\|SHAP value\|) | Variables | mean(\|SHAP value\|) |
| 1 | count_pocketparks_800 | 0.0107 | ints_500 | 0.0138 | ints_300 | 0.0116 |
| 2 | sumarea_pocketparks_800 | 0.0097 | count_pocketparks_800 | 0.0062 | count_pocketparks_800 | 0.0039 |
| 3 | ints_500 | 0.0047 | msavi_5yrs_greenest_100 | 0.0044 | msavi_5yrs_all_500 | 0.0039 |
| 4 | msavi_5yrs_all_500 | 0.0037 | ndvi_5yrs_greenest_100 | 0.0034 | sumarea_pocketparks_800 | 0.0036 |
| 5 | sumarea_pocketparks_300 | 0.0036 | built_isa_300 | 0.0032 | built_isa_500 | 0.0033 |
| 6 | ndvi_5yrs_greenest_100 | 0.0034 | msavi_5yrs_greenest_500 | 0.0031 | msavi_5yrs_all_300 | 0.0031 |
| 7 | age | 0.0030 | msavi_5yrs_greenest_300 | 0.0031 | ints_500 | 0.0030 |
| 8 | household1 | 0.0029 | msavi_5yrs_all_100 | 0.0031 | edu | 0.0025 |
| 9 | bool_pocketparks_300 | 0.0029 | ndvi_5yrs_all_100 | 0.0028 | ndvi_5yrs_all_100 | 0.0015 |
| 10 | ndvi_5yrs_all_500 | 0.0025 | household1 | 0.0028 | ints_100 | 0.0014 |
